# A PFAS-steroid axis is associated with lipid and bile acid metabolism in MASLD

**DOI:** 10.64898/2026.04.01.26350019

**Authors:** Pauli Tikka, Aidan McGlinchey, Sami F. Qadri, Ilia Evstafev, Alex M. Dickens, Hannele Yki-Järvinen, Tuulia Hyötyläinen, Matej Orešič

## Abstract

**Background & Aims:** Per- and polyfluoroalkyl substances (PFAS) are persistent endocrine-disrupting chemicals associated with metabolic dysfunction, including metabolic dysfunction-associated steatotic liver disease (MASLD). While PFAS perturb lipid and bile acid (BA) metabolism in a sex-specific manner, the underlying mechanisms remain unclear. We tested whether steroid hormones mediate PFAS-associated metabolic alterations.

**Methods:** In 104 patients with biopsy-characterized MASLD, we performed sex-stratified analyses applied liquid chromatography coupled to mass spectrometry (LC-MS) for chemical analysis, integrating circulating steroids, PFAS exposure, hepatic lipidomics and BA profiles.

**Results:** Steroid hormones were associated with MASLD severity in a sexually-dimorphic manner. Dihydrotestosterone showed consistent inverse associations with steatosis, fibrosis, necroinflammation and insulin resistance, particularly in females. PFAS exposure was associated with altered steroid profiles, predominantly indicating suppressed steroidogenesis in females. These PFAS-associated hormonal changes were linked to downstream alterations in hepatic lipids and BAs. Mediation analysis supported indirect effects of PFAS on metabolic pathways *via* steroids, including testosterone/epi-testosterone-mediated effects on ether phospholipids and estradiol-mediated effects on lithocholic acid. Females exhibited stronger PFAS-steroid-BA associations, whereas males showed weaker, lipid-centric effects.

**Conclusions:** PFAS exposure is associated with sex-specific disruption of steroid hormone pathways that may link environmental exposure to lipid and BA dysregulation in MASLD. These findings identify steroid hormones as potential key mediators of PFAS-associated metabolic dysfunction and highlight sex as a critical determinant in environmental liver disease.

## Introduction

Metabolic dysfunction-associated steatotic liver disease (MASLD) affects ∼30% of the global population and is characterized by hepatic lipid accumulation, insulin resistance, and progressive liver injury (Younossi et al. 2023). While obesity and metabolic syndrome are key drivers, environmental factors, particularly endocrine-disrupting chemicals (EDCs), are increasingly implicated in disease initiation and progression (Deierlein et al. 2017; Foulds et al. 2017; Giulivo et al. 2016; Heindel et al. 2017). EDCs can perturb endocrine signaling and metabolic pathways, acting either as early triggers or as accelerators of disease progression toward steatohepatitis and fibrosis (Treviño and Katz 2018; Vujkovic et al. 2022; Wahlang et al. 2019).

Per- and polyfluoroalkyl substances (PFAS) are ubiquitous, persistent EDCs associated with dysregulated lipid metabolism, bile acid (BA) homeostasis, and liver injury (Zhang et al. 2024). We previously demonstrated sex-specific metabolic effects of PFAS in MASLD, with stronger and predominantly adverse associations in females, including altered lipid and BA profiles (Sen et al. 2022). However, the mechanisms underlying this sexual dimorphism remain unresolved.

Steroid hormones are central regulators of hepatic metabolism, modulating lipid homeostasis, insulin sensitivity, and inflammation (Ortiz-Huidobro et al. 2021; Shen and Shi 2015; Song and Choi 2022; Yang et al. 2021). Glucocorticoids promote lipid accumulation and insulin resistance (Polyzos and Targher 2024), whereas estrogens generally exert protective metabolic effects (Yang et al. 2014). Androgens exhibit context-dependent and sex-specific actions, contributing to divergent metabolic phenotypes in males and females. Experimental and epidemiological evidence indicates that PFAS can disrupt steroidogenesis and hormone signaling (Cui et al. 2020; Joensen et al. 2013; Kang et al. 2016; Wang et al. 2021; Zhou et al. 2017), but whether these alterations associate with PFAS-linked metabolic dysfunction is unknown.

Here, we investigated steroid hormones as potential mediators of PFAS effects in MASLD. We examined (i) associations between steroids and disease severity, (ii) relationships between PFAS exposure and steroid profiles, and (iii) whether steroid alterations link PFAS exposure to changes in hepatic lipid and BA metabolism. By integrating clinical, metabolomic, and mediation analyses, we aimed to define potential key endocrine mechanisms underlying sex-specific environmental effects in MASLD.

## Methods

### Clinical cohort

The study population has been described previously (Sen et al. 2022). Briefly, 104 patients (69 females, 35 males) undergoing laparoscopic bariatric surgery were included (**Table 1**).

**Table 1.** The subject sizes in each case (steatosis, fibrosis, necroinflammation HOMA-IR) of this study. In addition to the overall case (HOMA-IR case level > 1.5 (Sen et al. 2022), and other cases (steatosis, fibrosis, necroinflammation) > 0) and control (≤ 1.5 or 0, respectively) group sizes, the study groups have been also subdivided to females and males.

|  | Steatosis |  |  | Fibrosis |  |  |
| --- | --- | --- | --- | --- | --- | --- |
|  | All | Females | Males | All | Females | Males |
| **Control** | 83 | 58 | 25 | 62 | 43 | 19 |
| **Case** | 21 | 11 | 10 | 42 | 26 | 16 |
|  | Necroinflammation |  |  | HOMA-IR |  |  |
|  | All | Females | Males | All | Females | Males |
| **Control** | 88 | 60 | 28 | 18 | 13 | 5 |
| **Case** | 16 | 9 | 7 | 86 | 56 | 30 |

Inclusion criteria were: (i) age 18–75 years; (ii) absence of acute or chronic diseases other than obesity, type 2 diabetes or hypertension; (iii) alcohol consumption <20 g/day (women) or <30 g/day (men); (iv) no evidence of other liver diseases or inborn metabolic disorders; and (v) no exposure to hepatotoxic drugs. Elevated liver enzymes were not an exclusion criteria. Clinical measurements were performed as described previously (Luukkonen et al. 2016). Demographic characteristics of the study population is shown in **Table S1**. Cases were defined as steatosis, fibrosis, or necroinflammation >0, and HOMA-IR >1.5, with corresponding controls defined as ≤0 or ≤1.5, respectively (Sen et al. 2022).

This is a cross-sectional observational study; therefore, causal inference cannot be established.

The study protocol was approved by the Ethical Review Committee of the Hospital District of Helsinki and Uusimaa (284/13/03/01/13). The study was conducted in accordance with the Declaration of Helsinki. Each participant provided written informed consent after being explained the nature and potential risks of the study.

### Steroid analysis

The steroids were extracted using a liquid-liquid extraction method. 20 µL of internal standards (**Table S2**) solution was added to 100 µL of each serum sample and extracted with with tert-butyl methyl ether. The samples were vortex mixed for 10 minutes and the upper layer was collected, evaporated to under nitrogen flow at 40°C and reconstituted in 50 µL of solvent (acetonitrile:water, 3:7, v/v). Matrix quality control samples were prepared from the pooled serum sample using the same preparation procedure as the test samples. Blanks were prepared in a similar manner, but with saline instead of serum. The extracts were then stored at −80 °C until analysis.

Calibration curves contained 11-Deoxycorticosterone (DOC), 11b-OH-Androstenedione (11b-OHA4), 11-Deoxycortisol (S), 11-Ketodihydrotestosterone (11-KDHT), 11-Ketotestosterone (11-KT), 17a-OH-Pregnenolone(17a-OHP5), 17a-OH-Progesterone(17aOH-P4), 11-Ketoandrostenedione (11-KA4), Aldosterone (A), Androstenedione (A4), Androsterone (AN), Corticosterone (B), Cortisol (F), Cortisone (E), Dehydroepiandrosterone (DHEA), Dihydrotestosterone (DHT), Estradiol (E2), Estrone (E1), Pregnenolone (P5), Progesterone (P4), Testosterone / Epi-testosterone (T/Epi-T) were prepared by mixing 20 µL of solvent (acetonitrile:water, 3:7, v/v), 40 µL of internal standard solution and 40 µL of corresponding working solution (**Table S3**). The final calibration curve levels were corresponding to the following concentration levels in serum: 5.00, 10.0, 20.0, 50.0, 100, 200, 500, 1000, 2000, 10000, 20000, 50000, 100000, 200000 pM. To control the calculated concentration during the analytical run, solvent control samples were prepared. The working solution with a concentration corresponding of 10000 pM in serum was prepared utilizing the procedure for preparing the calibration curve.

The samples were analyzed using ultra-high-performance liquid chromatography tandem mass spectrometry method (UHPLC-MS/MS) (**Table S4).** Briefly, the UHPLC system used in this work was an ExionLC system from AB Sciex LLC (Framingham, MA, USA). The system was equipped with an autosampler (maintained at 10 °C), two pumps and a column thermostat (maintained at 35 °C; **Table S5**). Separations were performed on a Kinetex Biphenyl 100 Å (2.1 mm × 100 mm, particle size 1.7 µm) by Phenomenex (Torrance, CA, USA). The mass spectrometer coupled to the UHPLC was a SCIEX Triple Quad™ 7500 instrument from AB Sciex LLC interfaced with OptiFlow Pro ion source (ESI) ion source. Analyses were performed in both negative and positive ion modes and Sciex OS (SCIEX OS 3.0.0.3339) was used for all data acquisition. Quality control was performed throughout the run with use of solvent control samples, matrix control samples and blanks.

### Metabolomic and PFAS analyses

Hepatic lipidomics and metabolite profiling were performed as described previously (Luukkonen et al. 2016; Sen et al. 2022), including lipidomics, polar and semi-polar metabolites, and targeted bile acid analysis. PFAS concentrations were measured from serum samples as described by Sen et al. (Sen et al. 2022). Combined summary datasets are provided in **Tables S6–S7**.

### Statistical analyses

Data were log2-transformed and autoscaled to zero mean and unit variance prior to further analysis. Associations were assessed using Spearman correlations and linear models, adjusted for age, BMI, and sex where appropriate. Chord plots were constructed using non-rejection rate (NRR) thresholds (0.2 overall; lower thresholds for sex-stratified analyses; **Figs. S1–S2**). The levels of high vs. low HOMA-IR levels (case vs. control) were chosen to be above or below 1.5, respectively (Sen et al. 2022). The other cases (steatosis, fibrosis, necroinflammation) were defined as those having values above 0.

### Causal mediation analysis

Causal mediation analysis (CMA) was performed using the R package *mediation* (Kurz 2023) to assess whether steroid hormones mediate associations between PFAS and lipid or bile acid outcomes. Models included (i) a mediator model (M) linking PFAS to steroids, and (ii) an outcome model (Y) linking PFAS and steroids to metabolic outcomes.

All models were adjusted for age, BMI, and sex (Walker 2024). The following quantities were estimated: average causal mediation effect (ACME; indirect effect), average direct effect (ADE), total effect, and proportion mediated. Analyses were performed using 100 simulations with default parameters using the *mediation* package.

## Results

### Steroid hormones showed large biological variation

Steroid concentrations showed inter-individual variability spanning several orders of magnitude (**Table S6**). Cortisone was the most abundant steroid in both sexes, followed by dehydroepiandrosterone and 11β-hydroxyandrostenedione. In males, testosterone/epitestosterone was among the most abundant steroids.

The cohort was characterized by severe obesity (mean BMI >47), a predominance of females (66%), and an age range of 29–67 years. Notably, 68% of females were below the median menopausal age in Finland.

### Steroids are associated with the staging of MASLD

We next investigated associations between steroid hormones and steatosis grade, fibrosis stage, necroinflammation, and HOMA-IR, both in the full cohort and in sex-stratified analyses, considering both case–control comparisons and continuous disease stages (**Figs. 1**, **S3–S9**).

**Fig. 1.**
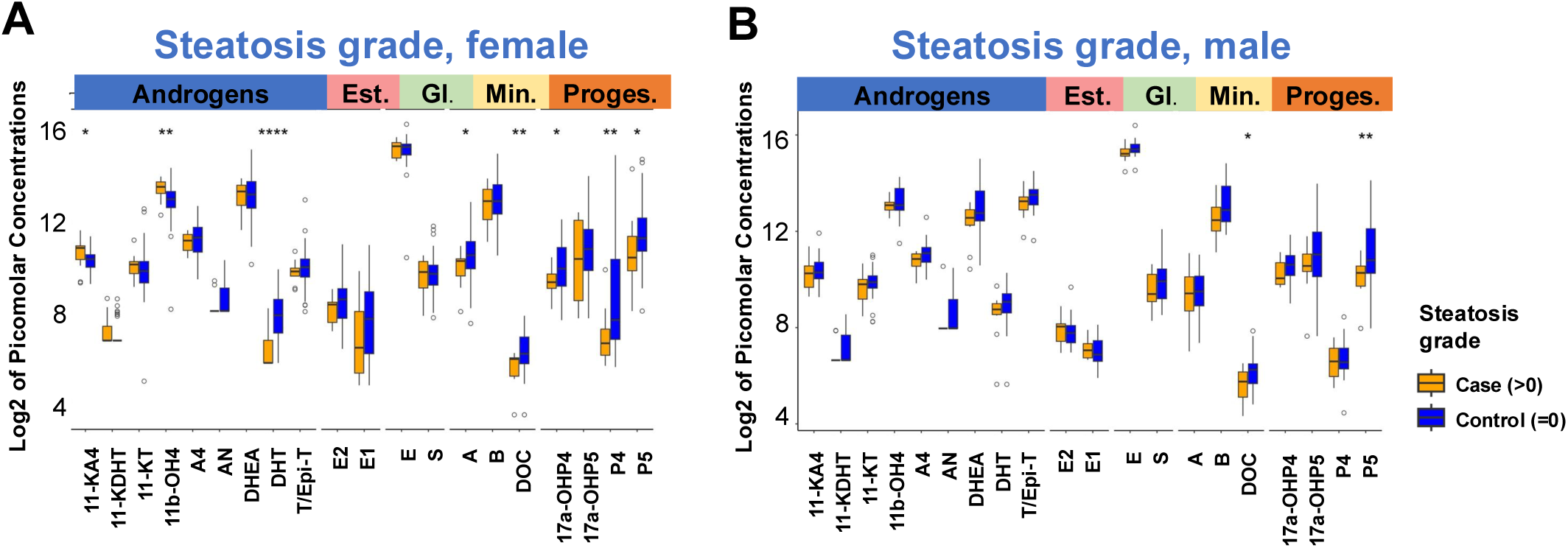
Steroid hormone concentrations in relation to steatosis. Log2-transformed plasma steroid levels in females (**A**) and males (**B**) stratified by steatosis status. Significant differences between groups are indicated (*p<0.1, **p<0.05, ***p<0.01, ****p<0.001; Wilcoxon test). The abbreviations of steroids: 11-Ketotestosterone (11-KT), 11b-OH-Androstenedione (11b-OH4), 11-Ketodihydrotestosterone (11-KDHT), 11-Ketoandrostenedione (11-KA4), Dehydroepiandrosterone (DHEA), Testosterone / (&) Epi-testosterone (T/Epi-T), Androsterone (AN), Dihydrotestosterone (DHT), Androstenedione (A4), Estradiol (E2), Estrone (E1), Cortisone (E), 11-Deoxycortisol (S), Corticosterone (B), Aldosterone (A), 17a-OH-Pregnenolone (17a-OHP5), 17a-OH-Progesterone (17a-OHP4), 11-Deoxycorticosterone (DOC), Pregnenolone (P5), Progesterone (P4), Estrogens (Est.), Glucocorticosteroids (Gl.), mineralcorticosteroids (Min.) and Progesterones (Proges.).

At the case–control level, steroid alterations were more pronounced in females than in males, particularly for steatosis (**Fig. 1A**). In the overall cohort, several steroids were lower in steatotic subjects (**Fig. S3**), including 11-deoxycortisol, pregnenolone, and progesterone. Pregnenolone and progesterone were also reduced in fibrosis, and progesterone was lower in necroinflammation. Subjects with fibrosis also showed reduced DHT levels.

In sex-stratified analyses, a broader pattern was observed in females, with eight steroids lower in steatosis (**Fig. S4**), including 11-ketoandrostenedione, 11β-hydroxyandrostenedione, DHT, aldosterone, 11-deoxycorticosterone, 17α-OH-progesterone, pregnenolone, and progesterone. In males, only 11-deoxycorticosterone and pregnenolone were significantly lower (**Fig. S5**). Similar patterns were observed across fibrosis, necroinflammation, and HOMA-IR (**Figs. S3–S5**).

Across continuous disease stages (**Figs. S6–S9**), DHT showed consistent negative associations with steatosis grade, fibrosis stage, necroinflammation, and HOMA-IR in the overall cohort. These associations were primarily driven by females, as DHT was significantly associated with these outcomes only in females in sex-stratified analyses.

Additional associations were observed for multiple steroids. Steatosis grade was negatively associated with 11-deoxycorticosterone, 11β-hydroxyandrostenedione, pregnenolone, and progesterone. Fibrosis stage was associated with testosterone/epitestosterone, while necroinflammation showed associations with androsterone, 11-deoxycortisol, corticosterone, pregnenolone, and progesterone. These patterns were generally consistent across sexes, although not all reached statistical significance in each subgroup.

HOMA-IR showed positive associations with 11β-hydroxyandrostenedione and 11-deoxycortisol in the overall cohort, and additionally with 11-ketotestosterone, 11-ketoandrostenedione, 11-deoxycortisol, and cortisone in males. Overall, androgens showed more associations with clinical parameters in males, predominantly negative, except for positive associations of 11-ketotestosterone and 11-ketoandrostenedione with HOMA-IR.

Age was negatively associated with several steroids (androstenedione, androsterone, dehydroepiandrosterone, estradiol, and progesterone) in the overall cohort, while 11β-hydroxyandrostenedione showed positive associations (**Figs. S10–S11**). Most of these associations were driven by females, except for dehydroepiandrosterone, which showed negative associations only in males. In males, androstenedione, estradiol, and progesterone were not associated with age.

BMI was associated with several steroids only in males, with positive associations observed for estradiol, estrone, androstenedione, and 11-ketotestosterone, and negative associations for androsterone and DHT.

### PFAS are associated with disease stage and steroid profiles in sex specific manner

We next examined associations between PFAS (**Table 7**), steroid hormones, and disease stage. PFAS exposure was associated with steroid profiles in a sex-specific manner (**Fig. 2**), with predominantly negative associations between PFAS and steroids. PFAS were also positively associated with HOMA-IR and age, particularly in females. Several steroids, including cortisone, corticosterone, aldosterone, 17α-OH-progesterone, and pregnenolone, were associated with multiple PFAS in the overall cohort.

**Fig. 2.**
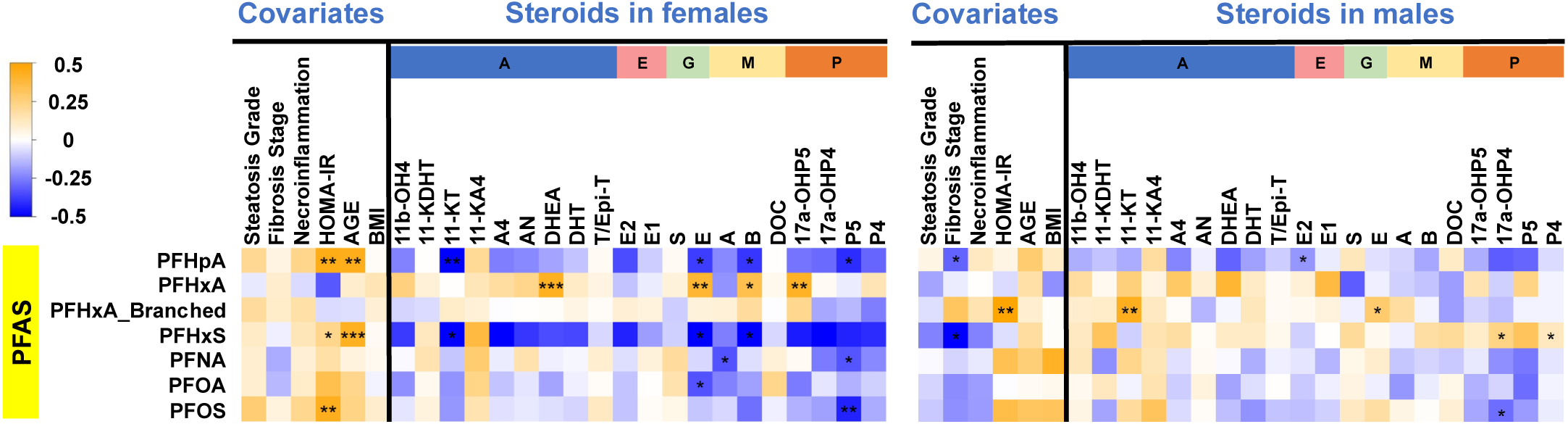
Associations between PFAS exposure, steroid hormones and clinical covariates. Linear model estimates are shown for females and males separately. Models were adjusted for age, BMI and sex where appropriate. Statistical significance is indicated (*p<0.05, **p<0.01, ***p<0.001).

PFHpA showed the strongest associations, with negative relationships to seven steroids, particularly progestogens, and these effects were most pronounced in females. PFNA and PFOS showed similar but weaker patterns, while PFOA showed a negative association with aldosterone.

PFHxS exhibited negative associations with several steroids in females (including 11-ketotestosterone, cortisone, and corticosterone), whereas in males it showed an opposite trend with positive associations for progestogens (17α-OH-progesterone and progesterone). In contrast, PFHxA showed predominantly positive associations with steroids, particularly dehydroepiandrosterone.

### Steroids are associated with bile acids and lipids

We next investigated associations between steroids and bile acids and lipids (**Table S7**). Steroid–metabolite associations showed clear sex-specific patterns (**Fig. 3**, **Fig. S12**).

**Fig. 3.**
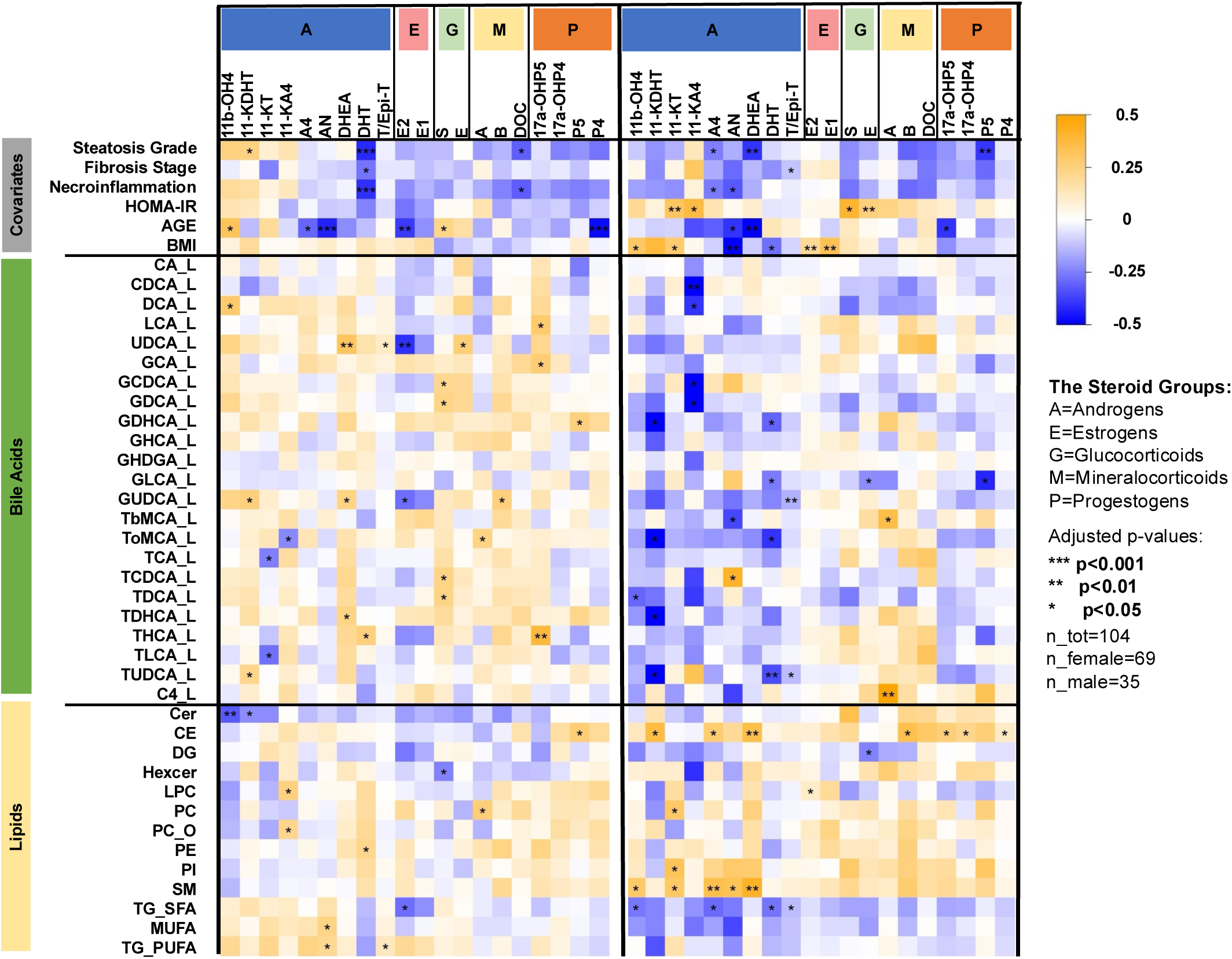
Associations between steroid hormones, bile acids and lipid classes. Heatmap of scaled linear model estimates (range −0.5 to 0.5) for associations between clinical variables, steroids, bile acids and lipid classes in females and males. Models adjusted for age, BMI and sex where appropriate.

When analyzing the data from both males and females simultaneously, steroids showed both positive and negative associations with bile acids and lipids, with positive associations largely driven by females. In females, many bile acids were positively associated with steroids, with the exception of selected estrogens and androgens. In contrast, in males, bile acids showed predominantly negative associations with steroids, particularly with androgens.

Associations between steroids and lipids were more similar across sexes but were generally stronger in males. In males, several steroids, particularly androgens and progestogens, were positively associated with cholesteryl esters, sphingomyelins, and phospholipids (PI, PC_O). Saturated triacylglycerols showed negative associations with androgens in males and with estradiol in females. In females, androgens showed negative associations with ceramides and positive associations with phospholipids (LPC, PC_O) and triacylglycerol groups (TG_SFA, TG_MUFA, TG_PUFA).

Network-level analysis (**Fig. 4**) revealed strong interconnections between PFAS, steroids, clinical variables, lipids, and bile acids. PFAS were closely linked to age and sex, and to several steroids. Lipid classes, particularly ceramides and diacylglycerols, showed correlations with both steroids and bile acids. Further analysis using network visualization (**Fig. S13**) showed that lipids clustered more closely with steroids, whereas bile acids formed a more distinct group with weaker direct associations with steroids.

**Fig. 4.**
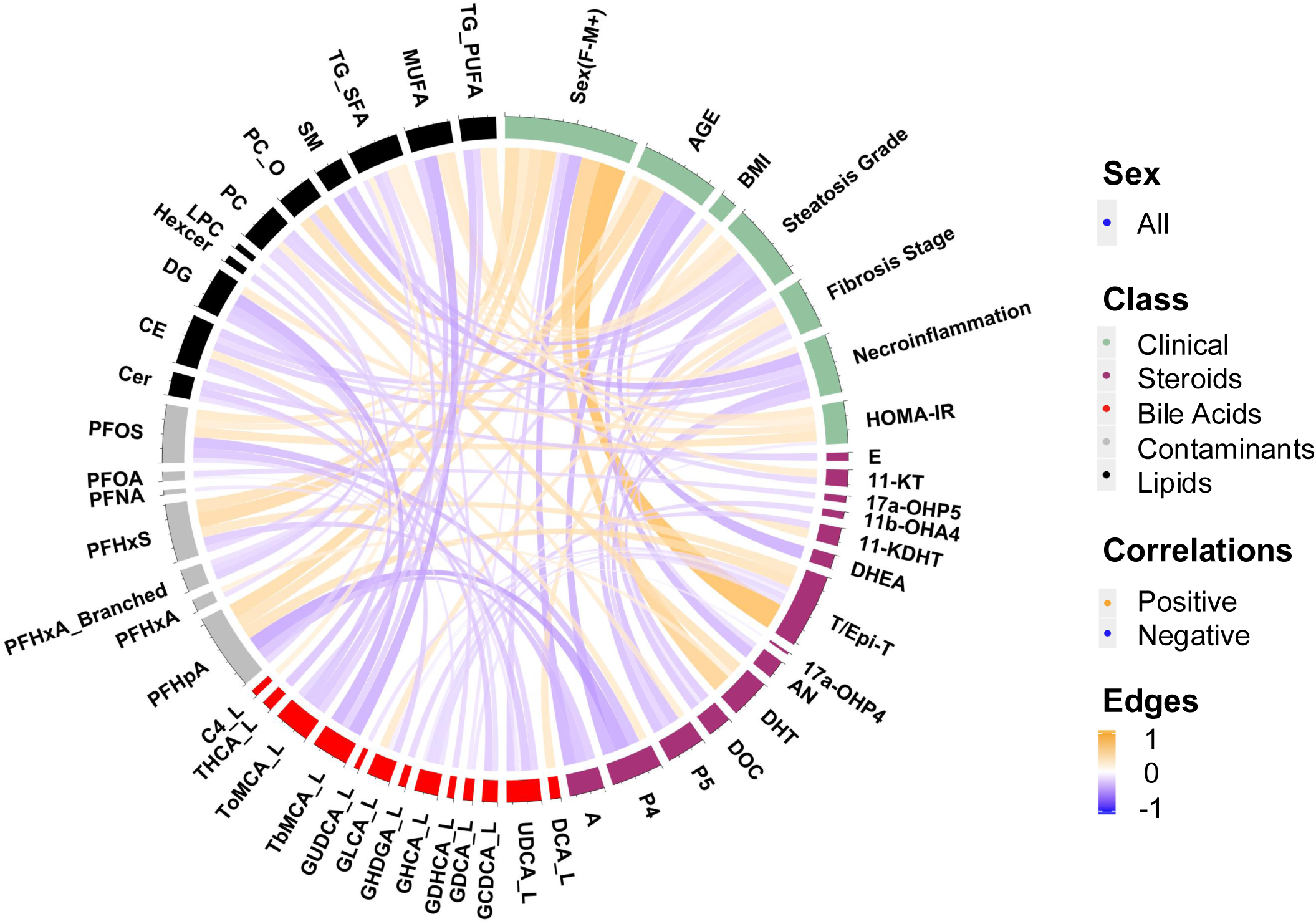
Network representation of associations between PFAS, steroids, bile acids, lipids and clinical variables. Chord diagram showing significant partial correlations (Spearman; NRR<0.03). Only strongest associations are displayed.

### Impacts of PFAS exposure on lipid metabolism may be mediated through steroids and bile acids

To assess whether steroid hormones may link PFAS exposure to metabolic outcomes, we performed causal mediation analysis (CMA). Indirect and direct effects for all subjects are shown in **Fig. 5** and **Table S8**. **Fig. 5A** illustrates the conceptual framework, including average causal mediation effects (ACME) and average direct effects (ADE), while **Fig. 5B** summarizes mediation pathways across all subjects.

**Fig. 5.**
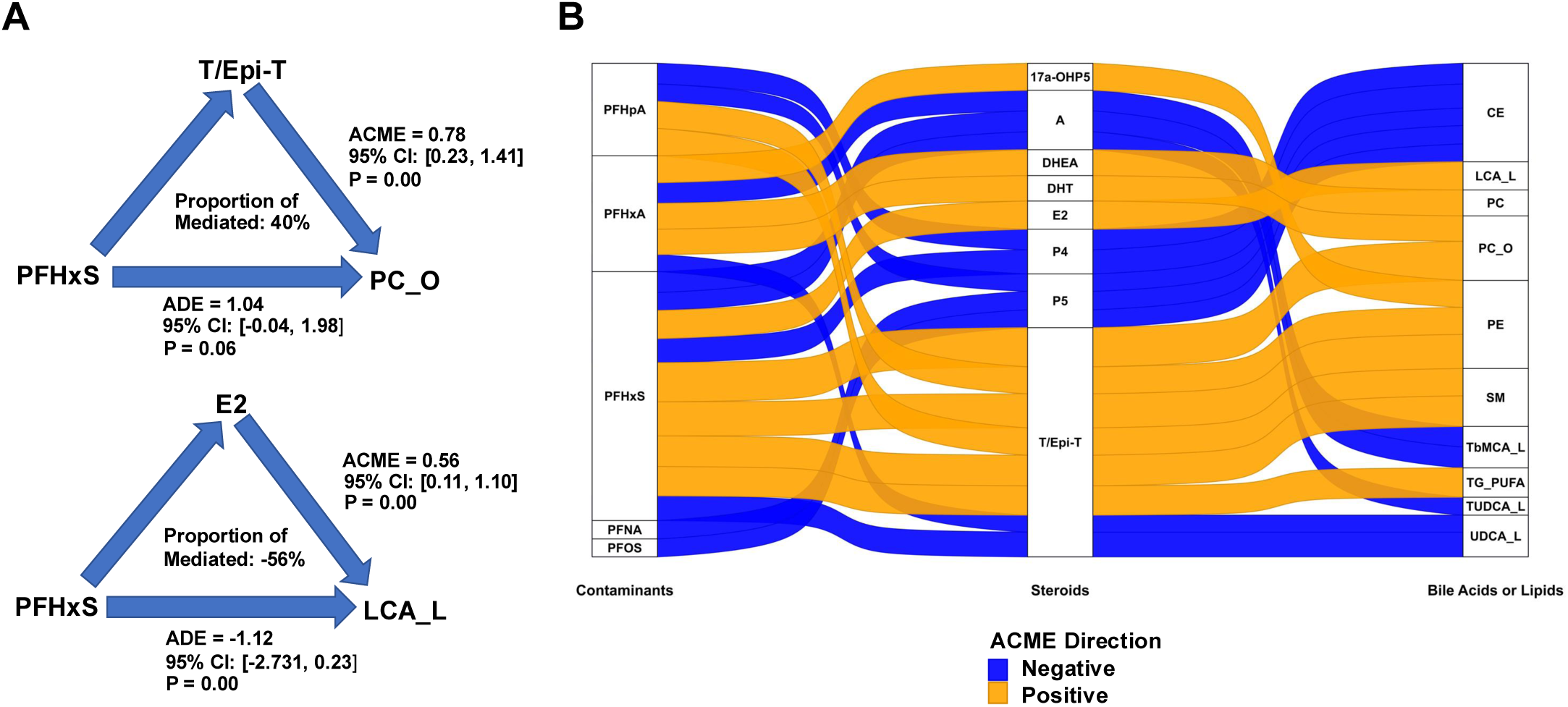
Causal mediation analysis of PFAS effects on metabolic pathways via steroid hormones. (**A**) Conceptual framework of mediation analysis, with two illustrative examples. (**B**) Sankey diagram illustrating indirect (ACME) and direct (ADE) effects of PFAS on lipid and bile acid outcomes mediated by steroid hormones. Only associations with p<0.1 are shown.

Potential steroid-mediated effects of PFAS on lipid and bile acid outcomes were suggested, with testosterone/epitestosterone (T/Epi-T) showing the most pronounced mediation effects (**Fig. 5B**). Sex-stratified analyses (**Figs. S14A–B**; **Tables S9–S10**) identified different dominant mediation effects: in females, dehydroepiandrosterone (DHEA) and 17α-OH-pregnenolone were the primary mediators, whereas in males 11-ketotestosterone and pregnenolone were most prominent. These sex-specific mediation patterns were not altered after adjustment for age and BMI (**Fig. S15**).

## Discussion

Steroid hormones are central regulators of hepatic lipid metabolism, insulin sensitivity and inflammation (Dobre et al. 2025; MacMillan et al. 2024; Quagliarini et al. 2023). In this study, we find that a sex-dependent network may link steroid hormones to metabolic dysfunction-associated steatotic liver disease (MASLD) severity, PFAS exposure, and downstream lipid and bile acid (BA) metabolism (Henriksen et al. 2016; India-Aldana et al. 2023; Lee et al. 2011; Seidemann et al. 2024).

Importantly, this study extends prior PFAS research by demonstrating that steroid hormones are not only associated with PFAS exposure but may act as intermediate links between PFAS and hepatic metabolic alterations. While previous studies have reported associations between PFAS and lipid metabolism or hormone levels separately (India-Aldana et al. 2023; Sen et al. 2022), our integrative analyses suggest a connected PFAS–steroid–metabolism axis in MASLD.

DHT) showed the most consistent associations, particularly in females, across steatosis, fibrosis, necroinflammation and insulin resistance. DHT enhances insulin sensitivity and glucose uptake (Frank et al. 2019; Mårin 1995; Saad et al. 2001), yet its metabolic effects are context- and sex-dependent. Elevated androgen activity in females has been linked to insulin resistance and adverse metabolic phenotypes (Münzker et al. 2015; Münzker et al. 2017). This divergence is supported by experimental models showing opposing androgen effects on hepatic metabolism between sexes (Cherubini et al. 2024; Lin et al. 2008).

Dehydroepiandrosterone (DHEA) further highlighted sex-specific metabolic patterns. In males, DHEA was associated with cholesteryl esters and sphingomyelins—lipid classes implicated in MASLD progression (Ramos-Molina et al. 2024; Syed-Abdul 2023). In females, DHEA was instead associated with secondary BAs, suggesting involvement of gut microbiota in steroid metabolism. Microbial regulation of steroid conversion, including androgen metabolism, supports the presence of a gut–liver–steroid axis (Mayneris-Perxachs et al. 2020).

Several additional steroids, including progesterone and pregnenolone, were associated with disease severity and metabolic pathways, consistent with their established roles in hepatic lipid metabolism and insulin sensitivity (Yang et al. 2017). Notably, some steroids were linked to lipid and BA profiles independent of direct associations with disease stage, suggesting broader regulatory roles in hepatic metabolism.

PFAS exposure was associated with altered steroid hormone profiles in a sex-dependent manner, in line with previous evidence of PFAS-induced disruption of steroidogenesis (India-Aldana et al. 2023; Kushnir et al. 2024). In females, most associations were negative, including those involving pregnenolone, the precursor of all steroid hormones, suggesting reduced steroidogenic activity. Experimental studies indicate that PFAS can alter steroidogenic enzyme expression and hormone synthesis (Eggert et al. 2019; Tian et al. 2019). In contrast, associations in males were fewer and often directionally-opposite, although this may partly reflect smaller sample size.

While the cross-sectional design precludes causal inference, the integration of mediation analysis with multi-omics profiling supports a model in which steroid hormones may contribute to linking PFAS exposure to downstream metabolic alterations. In females, PFAS-associated steroids were primarily linked to BA metabolism, particularly secondary BAs, whereas in males associations were more lipid-centered. These findings suggest sex-specific metabolic routing of PFAS effects, potentially involving microbiota-dependent mechanisms.

Mediation analysis further supported indirect effects of PFAS on metabolic pathways via steroids. Estradiol mediated associations with lithocholic acid, a hepatotoxic BA linked to disease severity (Sen et al. 2024), whereas testosterone/epi-testosterone mediated associations with phospholipids previously associated with more favourable metabolic profiles (McGlinchey et al. 2022). These findings are consistent with our prior observations of stronger PFAS-associated metabolic disturbances in females (Sen et al. 2022).

Strengths of this study include comprehensive integration of clinical phenotyping with steroid profiling, lipidomics and BA analysis in a biopsy-characterised MASLD cohort. Limitations include the cross-sectional design and lack of detailed hormonal status information, particularly in women, which may influence steroid variability. In addition, PFAS and steroid measurements were obtained from serum, whereas lipid and BA profiles were derived from liver tissue.

In conclusion, our findings identify steroid hormones as potential key intermediates linking PFAS exposure to metabolic dysregulation in MASLD, with pronounced sex-specific patterns. These results provide a mechanistic framework for understanding environmental contributions to MASLD and highlight the importance of incorporating sex as a biological variable in both mechanistic studies and environmental health risk assessment.

## Declaration of interest

The authors declare that no competing interests exist.

## Financial support statement

This study was supported by the “Investigation of endocrine-disrupting chemicals as contributors to progression of metabolic dysfunction-associated steatotic liver disease” (EDC-MASLD) consortium funded by the Horizon Europe Program of the European Union under Grant Agreement 101136259 (to T.H. and M.O.), the Novo Nordisk Foundation (grant no. NNF20OC0063971 to T.H. and M.O.), Academy of Finland (grant no. 333981 to M.O. and no. 309263 to H.Y.-J.), Swedish Research Council (grant no. 2016-05176 to T.H and M.O), Juselius Foundation (to H.Y.-J.), the EPoS (Elucidating Pathways of Steatohepatitis) project funded by the Horizon 2020 Framework Program of the European Union (grant no. 634413, to M.O., H.Y.-J., and T.H.), the Swedish Knowledge Foundation (grant no. 20220122, to T.H. and M.O.), the Orion Research Foundation (to S.F.Q.), the Yrjö Jahnsson Foundation (20207313 to S.F.Q.), the Maud Kuistila Memorial Foundation (2021-0301B to S.F.Q.), the Emil Aaltonen Foundation (210182 to S.F.Q.), the Finnish Medical Foundation (5843 to S.F.Q.), and the Biomedicum Helsinki Foundation (20230241 to S.F.Q.). The authors would also like to thank the Turku Metabolomics Centre and Biocentre Finland for funding the instruments at the University of Turku used in this manuscript. Views and opinions expressed are those of the authors only and do not necessarily reflect those of the European Union. Neither the European Union nor the granting authority can be held responsible for them.

## Authors contributions

Conceptualization and design of study (MO, HYJ, TH), clinical study (SFQ, coordinated by HYJ), metabolomics analysis (IE, AMD, TH), data analysis and interpretation (PT, with critical input from SFQ, AM, HYJ, TH, supervised by MO), manuscript preparation (PT, MO, TH), critical review and editing (all authors).

## Data availability statement

Data from the clinical study are available upon request and an appropriate institutional collaboration agreement.

## Abbreviations

Bile acids
BPA: bisphenol A
CA: cholic acid
CErs: ceramides
CYP7A1: cholesterol 7a-hydroxylase
DCA: deoxycholic acid
DG: diacylglycerol
EC: environmental contaminants
EDCs: endocrine-disrupting chemicals
GCA: glycocholic acid
GCDCA: glycochenodeoxycholic acid
HCA: hyocholic acid
HDCA: hyodeoxycholic acid
HexCers: hexosylceramides
HOMA-IR: homeostatic model assessment of insulin resistance
hPPARa: humanized PPARa
IR: insulin resistance
LCA: lithocholic acid
LPCs: lysophosphatidylcholines
LPEs: lysophosphatidylethanolamines
MASLD: metabolic-dysfunction associated steatotic liver disease
MASH: metabolic-dysfunction associated steatohepatitis
PCs: phosphatidylcholines
PEs: phosphatidylethanolamines
PFAS: perfluorinated alkyl substances
PFHxS: perfluorohexanesulfonic acid
PFNA: perfluorononanoic acid
PFOA: perfluorooctanoic acid
PFOS: perfluorooctanesulfonic acid
SMs: sphingomyelins
TCA: taurocholic acid
TCDCA: taurochenodeoxycholic acid
TG: triacylglycerol
11-KT: 11-Ketotestosterone
11b-OHA4: 11b-OH-Androstenedione
11-KDHT: 11-Ketodihydrotestosterone
11-KA4: 11-Ketoandrostenedione
DHEA: Dehydroepiandrosterone
T/Epi-T: Testosterone / (&) Epi-testosterone
AN: Androsterone
DHT: Dihydrotestosterone
A4: Androstenedione
E2: Estradiol
E1: Estrone
E: Cortisone
S: 11-Deoxycortisol
B: Corticosterone
A: Aldosterone
17a-OHP5: 17a-OH-Pregnenolone
17a-OHP4: 17a-OH-Progesterone
DOC: 11-Deoxycorticosterone
P5: Pregnenolone
P4: Progesterone
A: Androgenes
€: Estrogens
G: Glucocorticosteroids
M: mineralcorticosteroids
P: Progesterones

## Supplementary figures

**Fig. S1.**
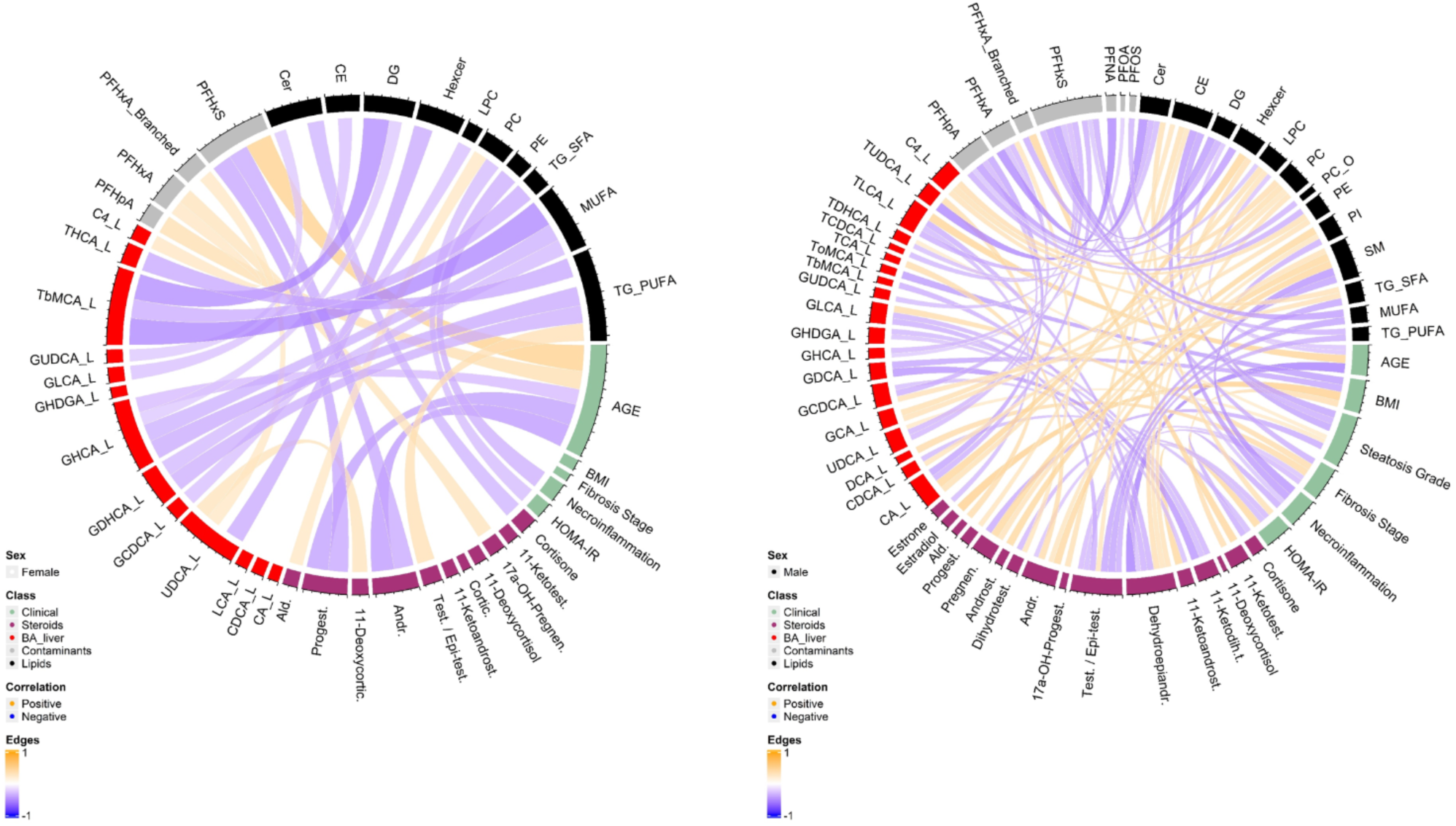
**Chord plot with significantly correlated (NRR<0.01) data of female and male subjects.**

**Fig. S2.**
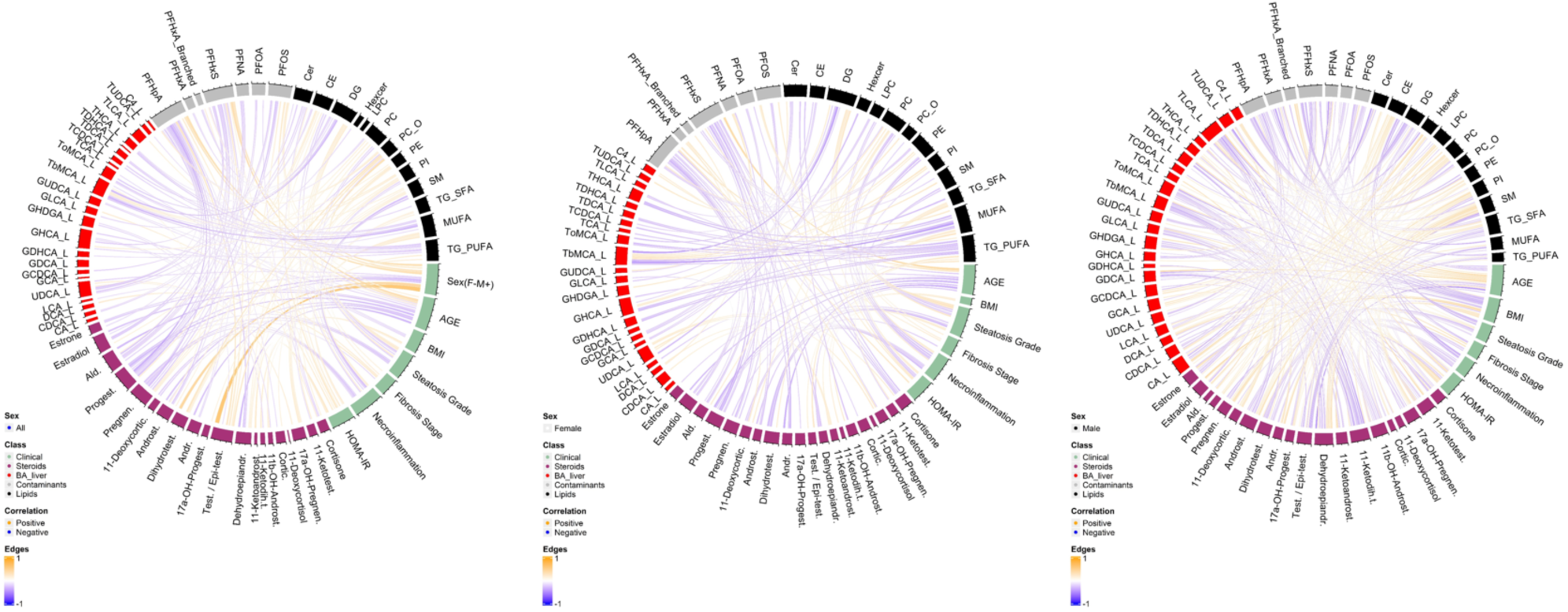
**A chord plot with significantly correlated (NRR<0.2) data of all, female and male subjects.**

**Fig. S3.**
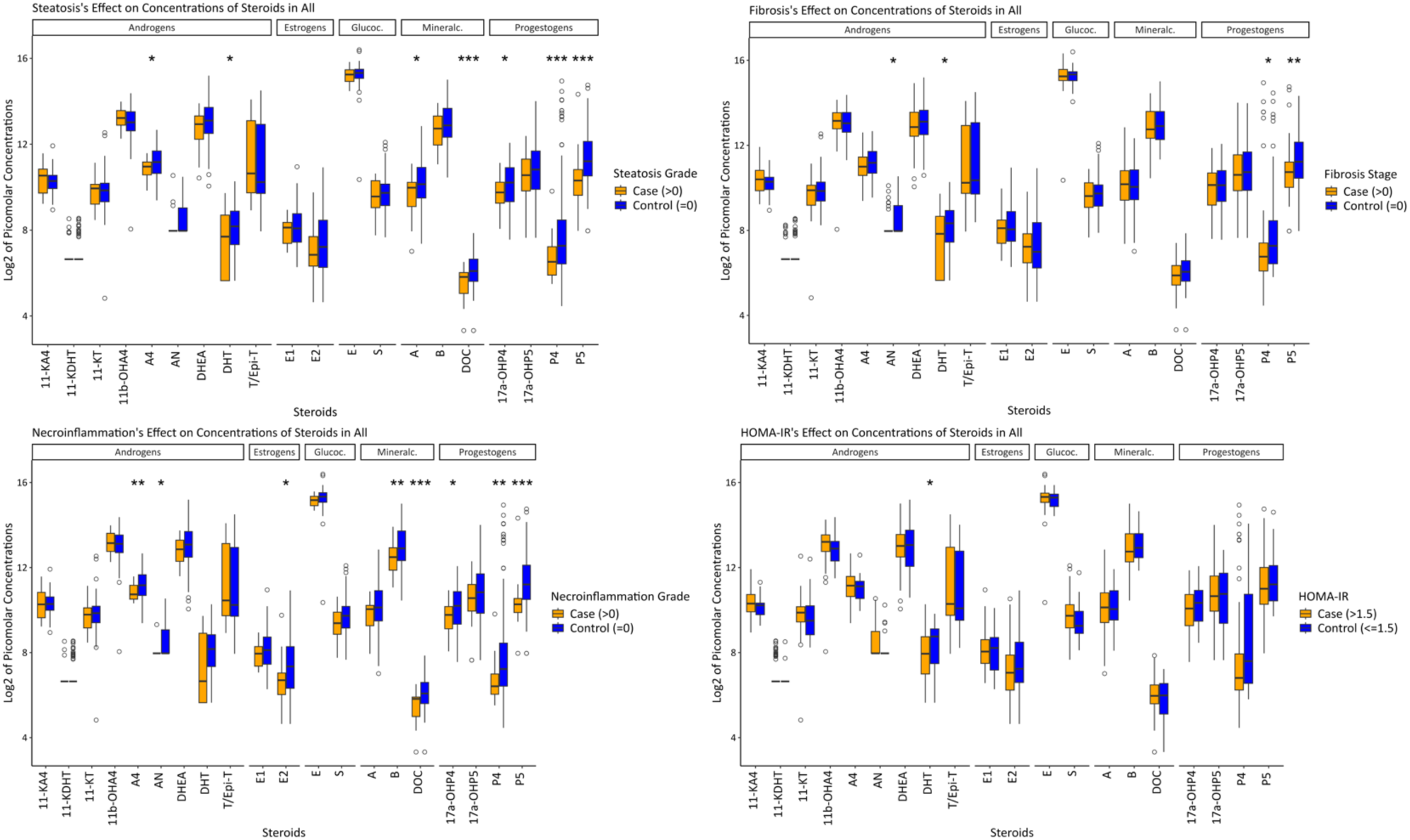
Log2 of the picomolar concentrations of steroids in the all subjects of the cases (Steatosis, Necroinflammation, Fibrosis, HOMA-IR). The cases (orange) and controls (blue) are defined at Fig. 1. The significant steroids as per the linear models are marked with an asterisk (*).

**Fig. S4.**
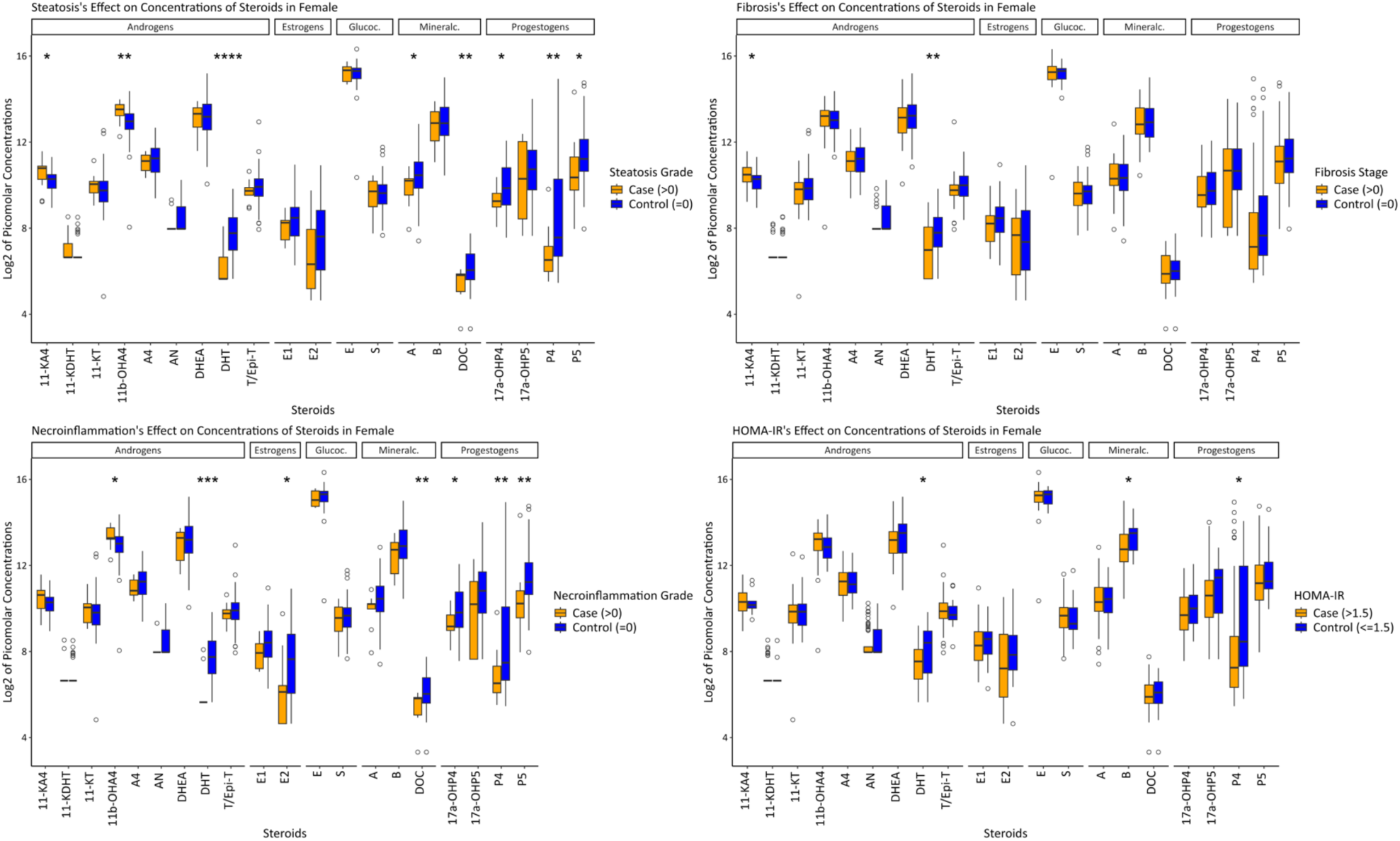
Log2 of the picomolar concentrations of steroids in the female subjects of the cases (Steatosis, Necroinflammation, Fibrosis, HOMA-IR). The cases (orange) and controls (blue) are defined at Fig. 1. The significant steroids as per the linear models are marked with an asterisk (*).

**Fig. S5.**
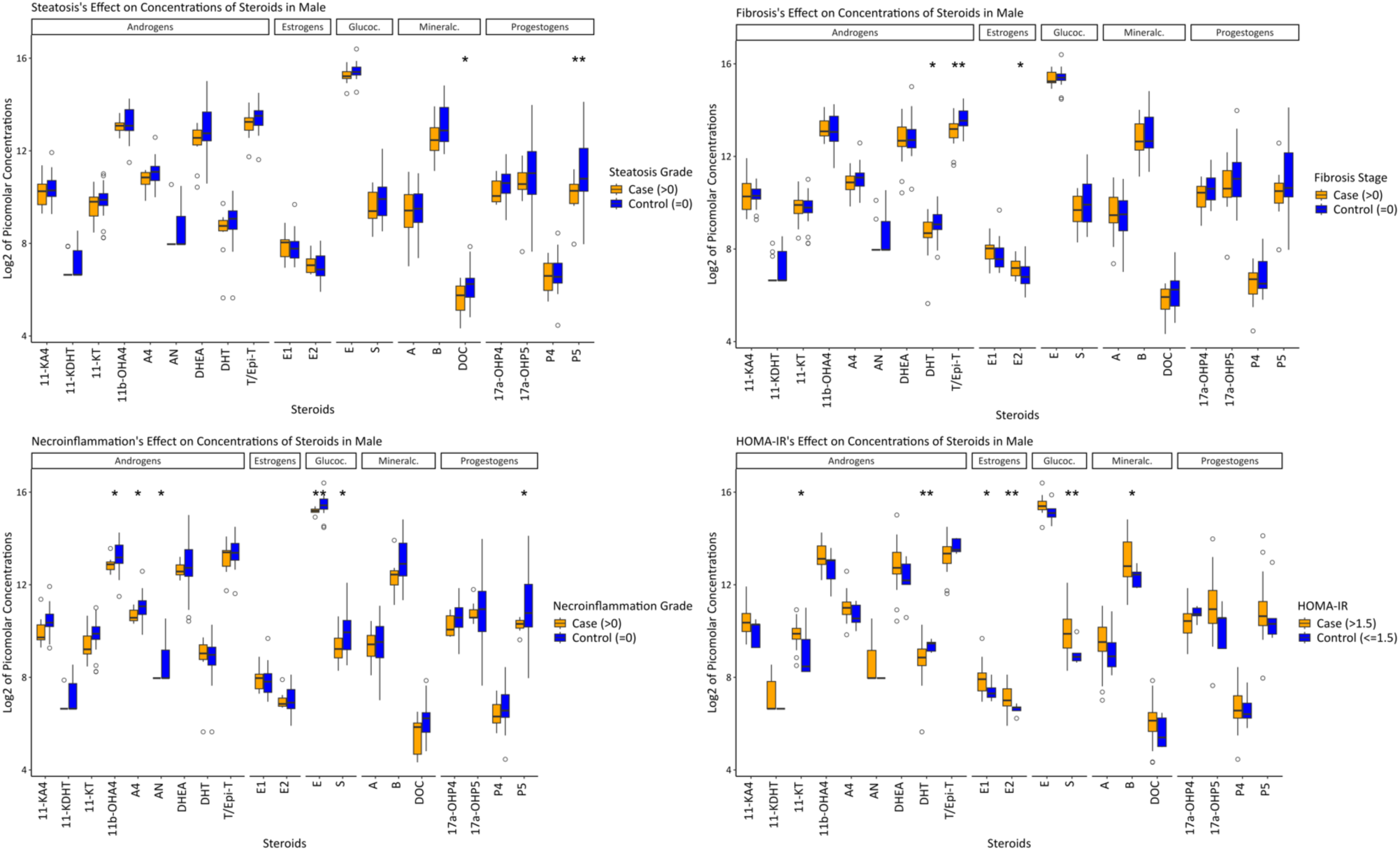
Log2 of the picomolar concentrations of steroids in the male subjects of the cases (Steatosis, Necroinflammation, Fibrosis, HOMA-IR). The cases (orange) and controls (blue) are defined at Fig. 1. The significant steroids as per the linear models are marked with an asterisk (*).

**Fig. S6.**
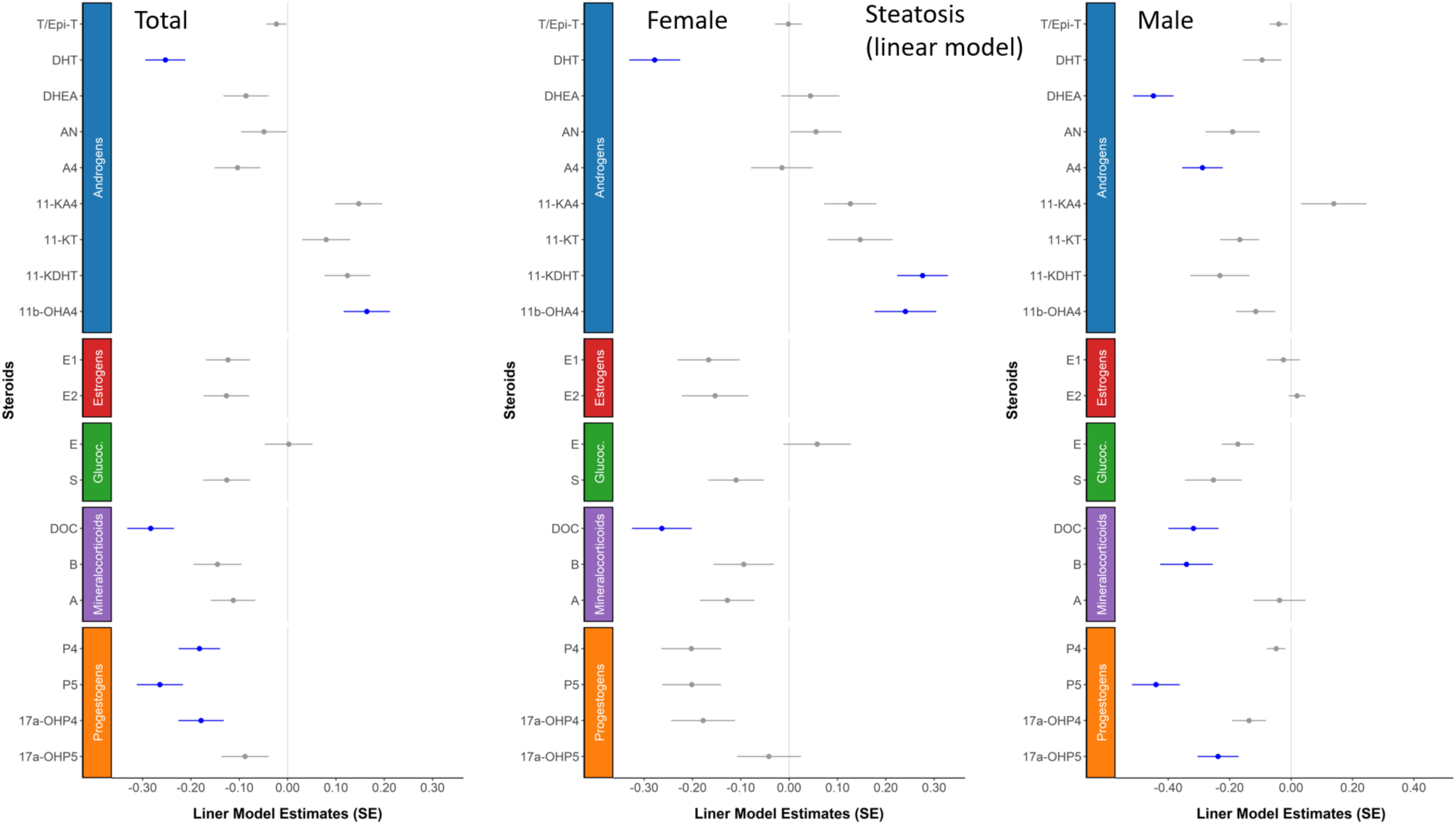
The association of steatosis on steroids. The data was shown in three groups: all subjects, female and male subjects. Linear model coefficients and the adjusted significances (Age, BMI, gender) were derived with MaasLin2 R package (**Table S11**).

**Fig. S7.**
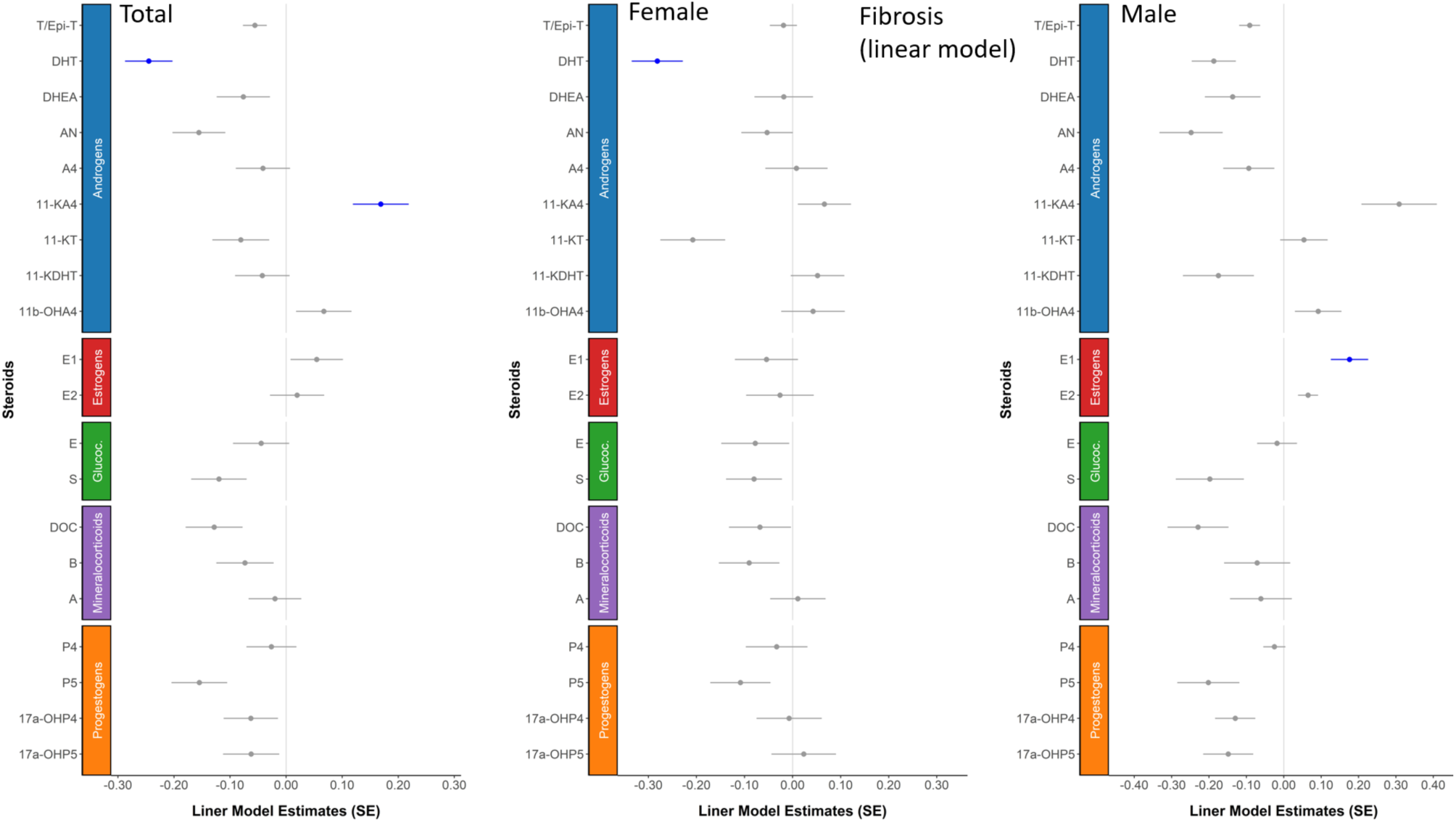
The association of fibrosis on steroids. The data was shown in three groups: all subjects, female and male subjects. Linear model coefficients and the adjusted significances (Age, BMI, gender) were derived with MaasLin2 R package (**Table S11**).

**Fig. S8.**
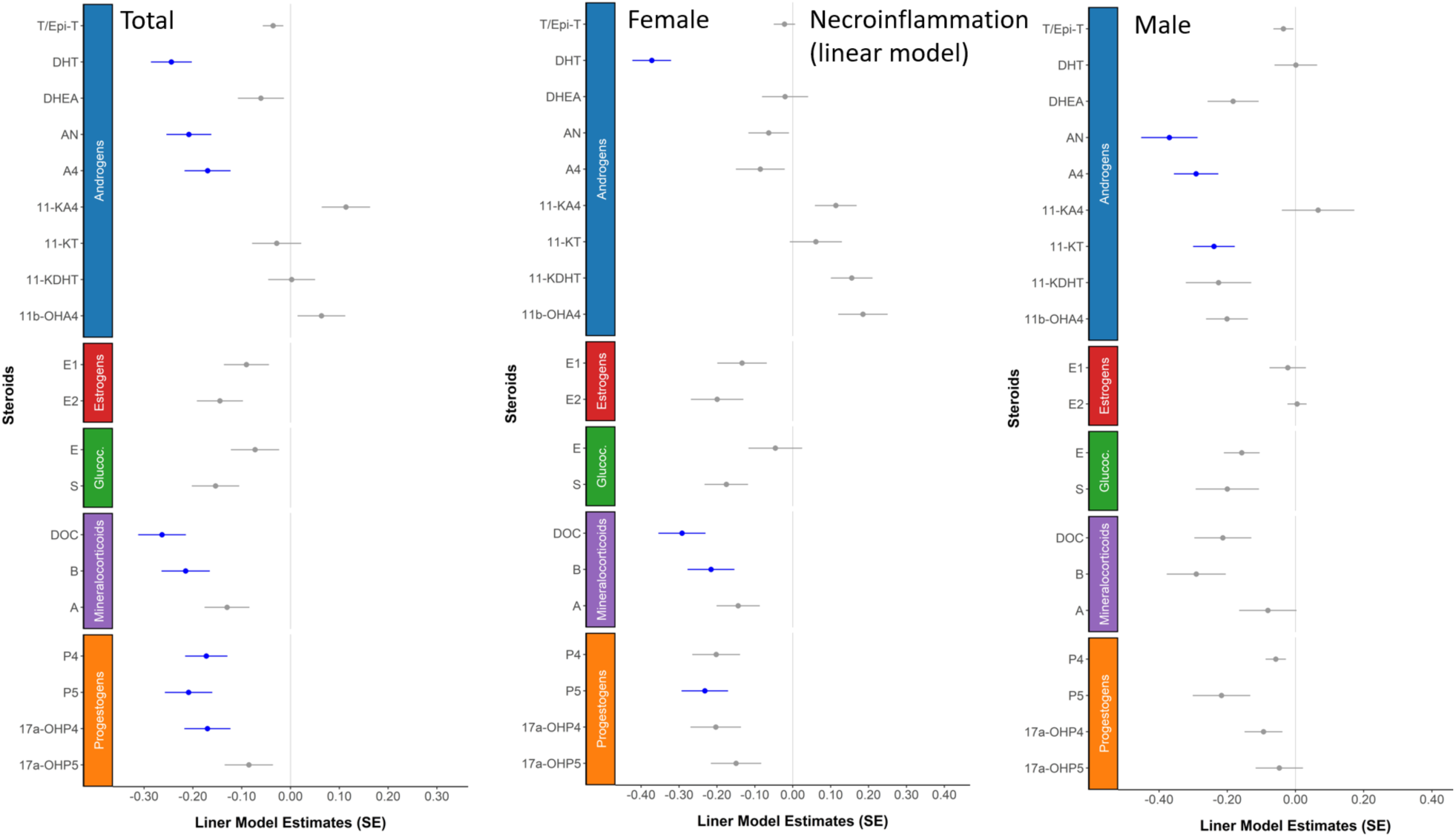
The association of necroinflammation on steroids. The data was shown in three groups: all subjects, female and male subjects. Linear model coefficients and the adjusted significances (Age, BMI, gender) were derived with MaasLin2 R package (**Table S11**).

**Fig. S9.**
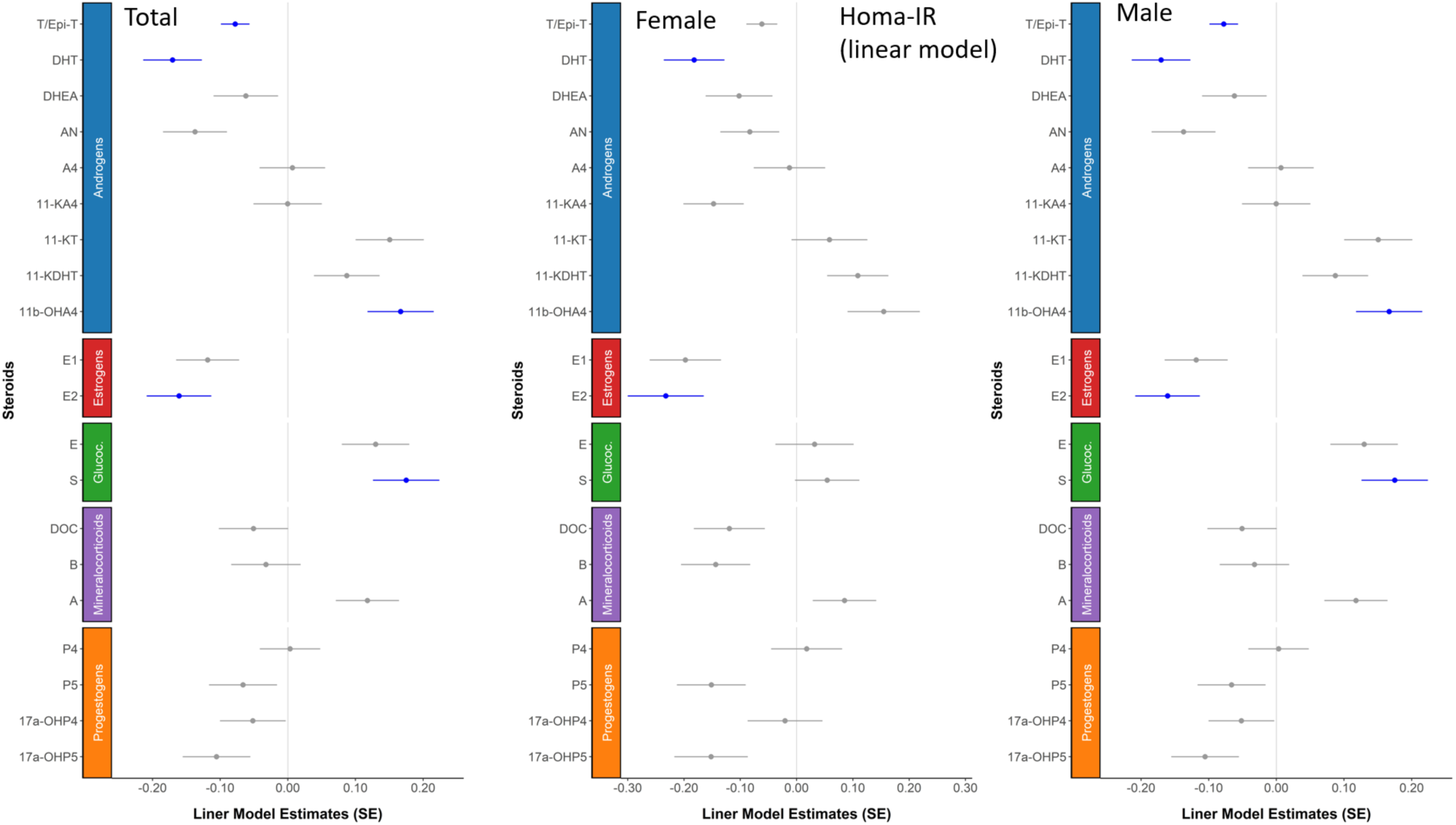
The association of HOMA-IR on steroids. The data was shown in three groups: all subjects, female and male subjects. Linear model coefficients and the adjusted significances (Age, BMI, gender) were derived with MaasLin2 R package (**Table S11**).

**Figure S10.**
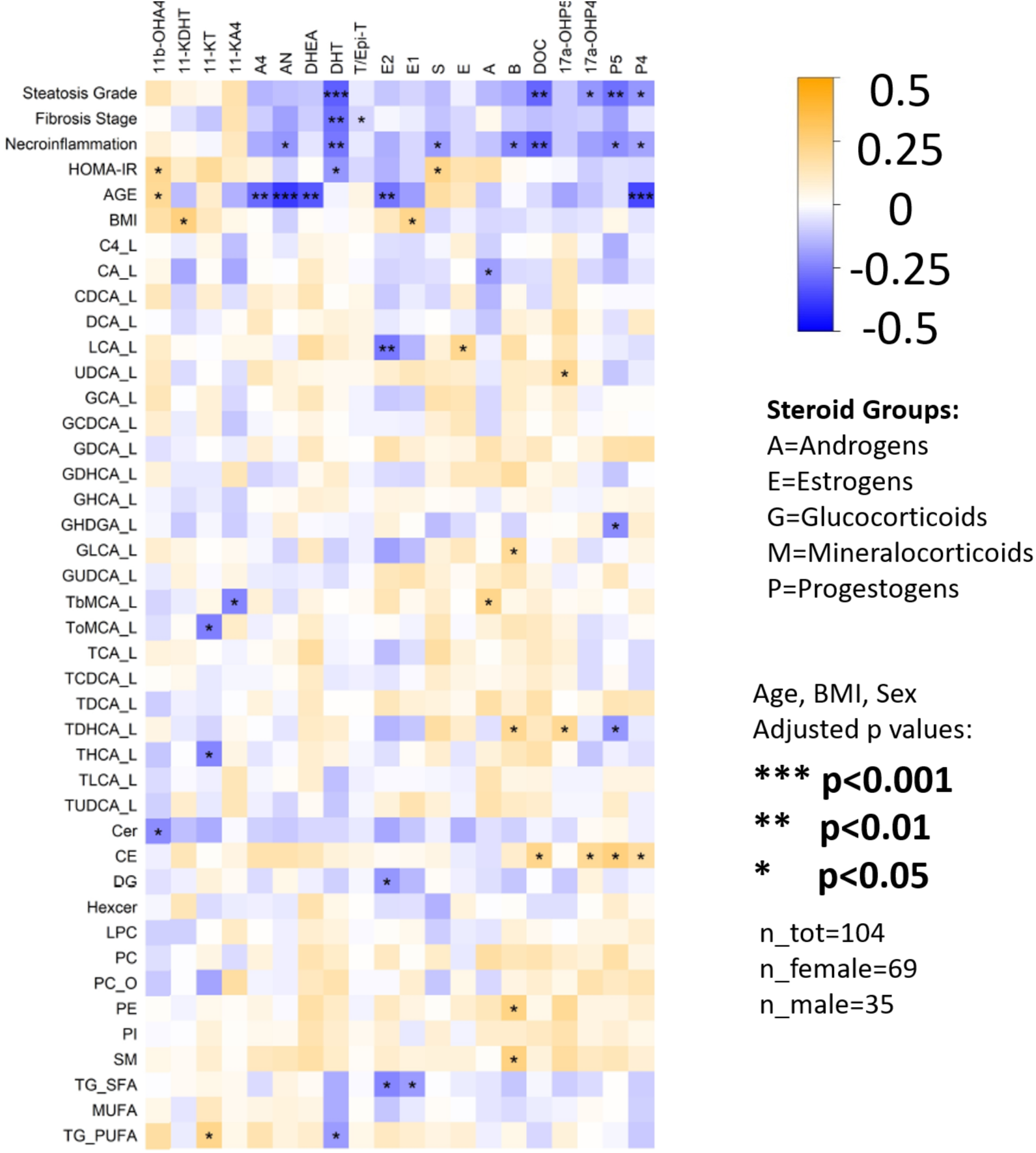
Linear model estimates between clinical covariates, bile acids, lipids and steroids in all subjects. The estimates were scaled between a range (−0.5 to 0.5) and the blue is a negative and red is a positive association. The confounders (age, BMI and sex) were added to the models when applicable. The p-values were obtained from the summary function (in R) with the models (via t tests; ***p<0.001, **p<0.01, *p<0.05). Steroid abbreviations are as in **Fig. 1**.

**Fig. S11.**
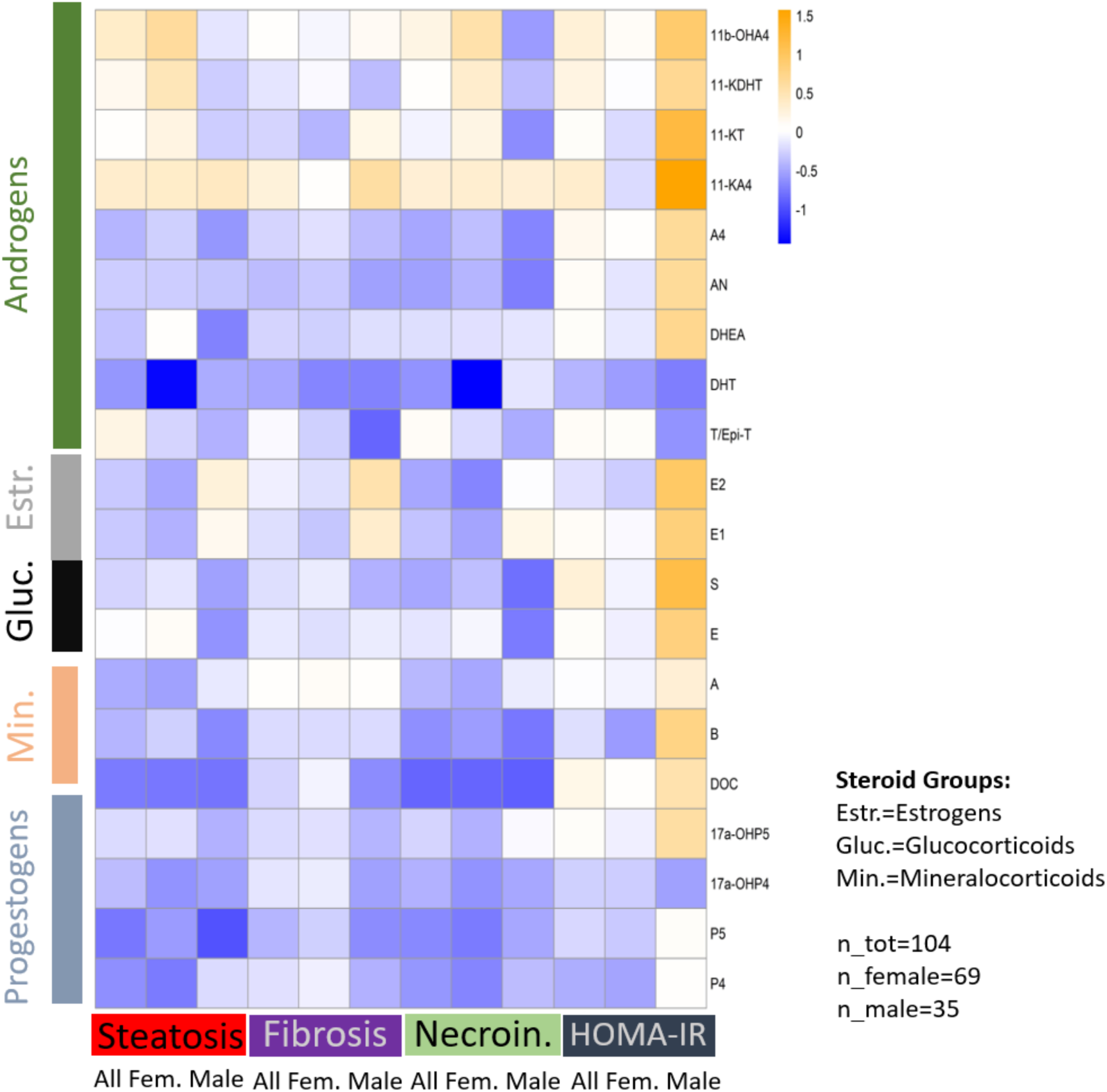
**Effect sizes in each group and sex.**

**Fig. S12.**
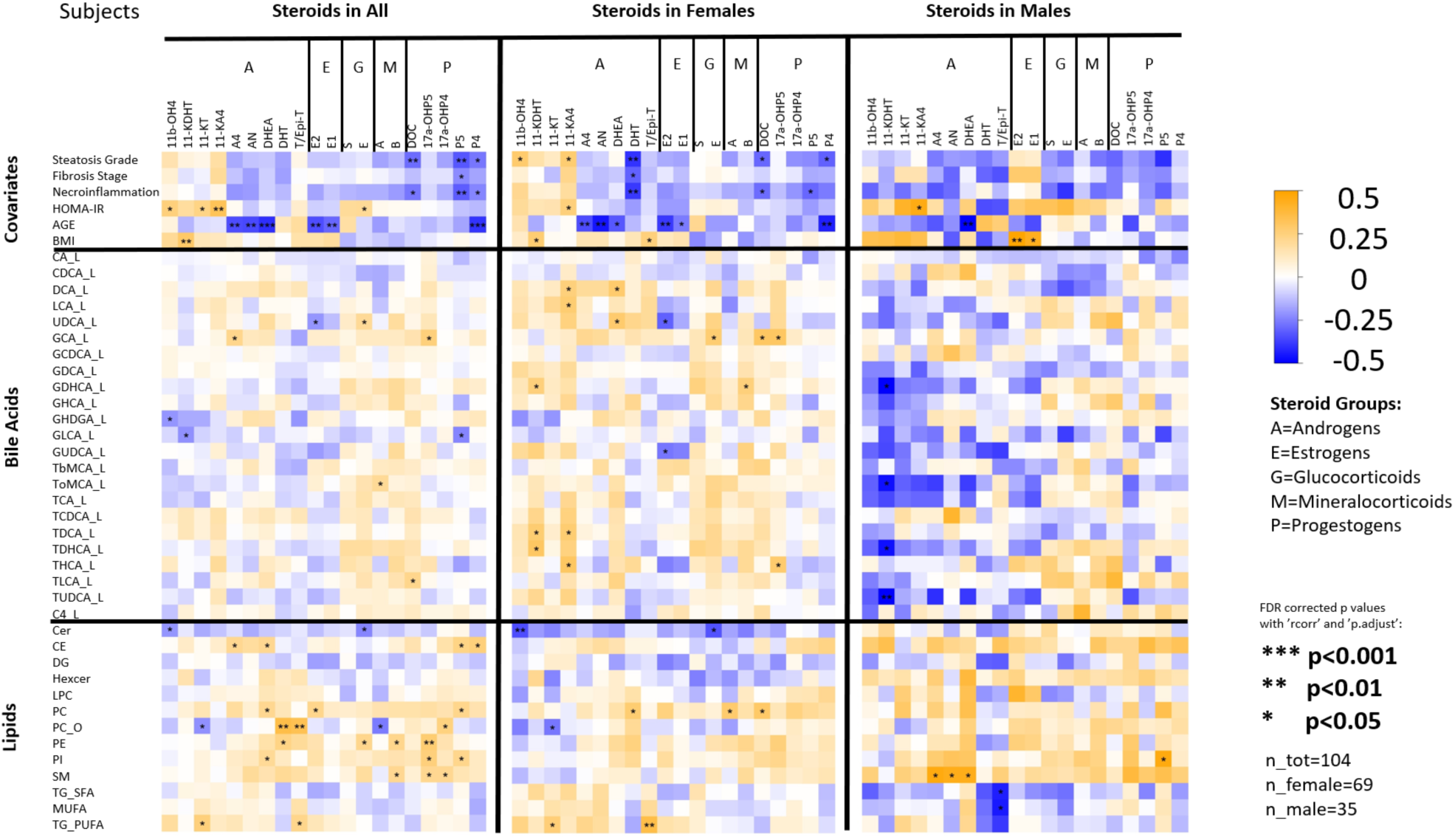
Correlation coefficients (Spearman) between bile acids, lipids, clinical covariates and steroids. The coefficients were scaled between a range (−0.5 to 0.5) for a visual comparison and the blue is a negative and red is a positive association. Panels were done with the corrplot function (in R). The p values have been corrected for multiple testing. **\*\*\*p<0.001, **p<0.05, *p<0.2.**

**Fig. S13.**
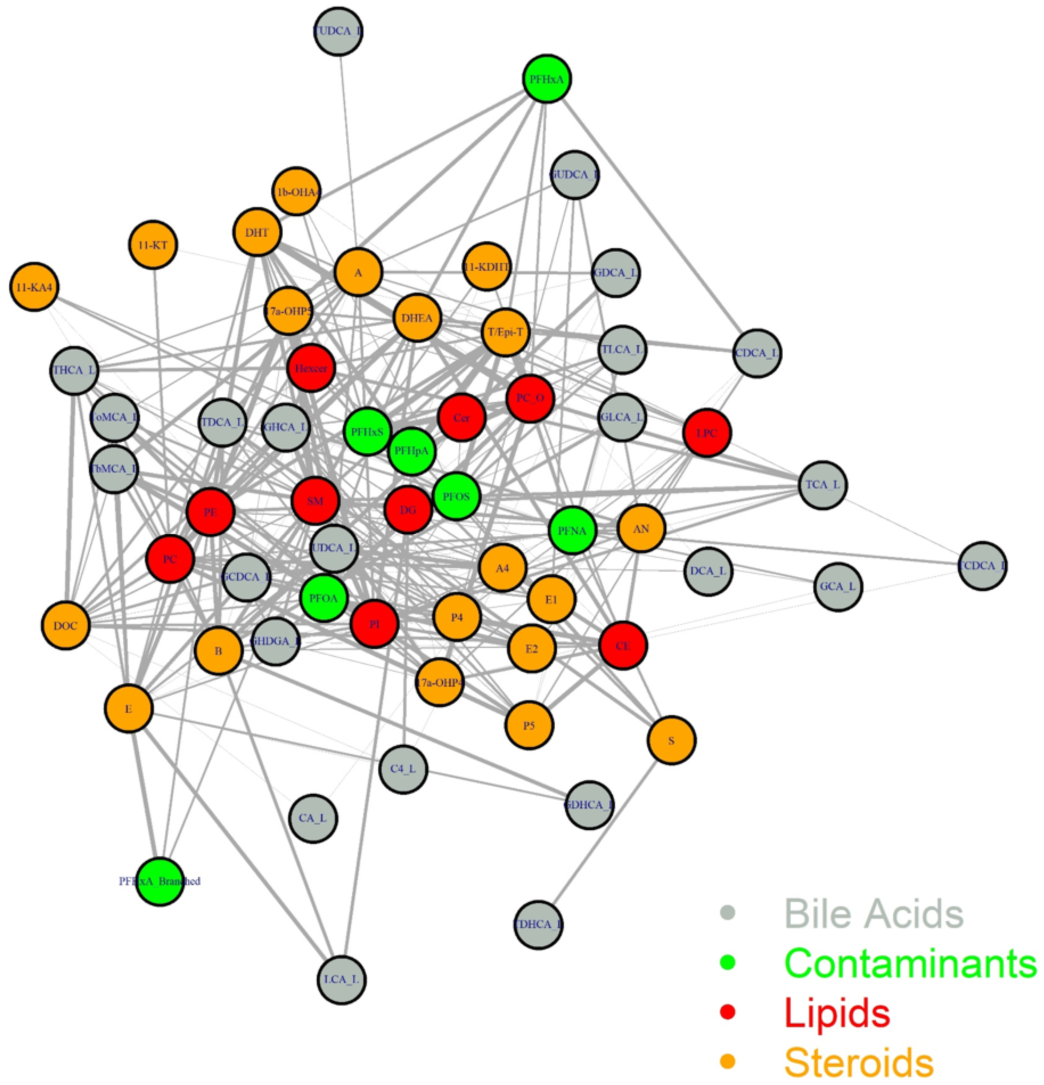
Network plot done with iGraph and network functions (in R) using all samples’ correlations with NRR=0.3 cut-off. The sizes of edges correspond the relative correlation strengths between the vertices. The vertex size is 0.3 times the degree of the network.

**Fig. S14.**
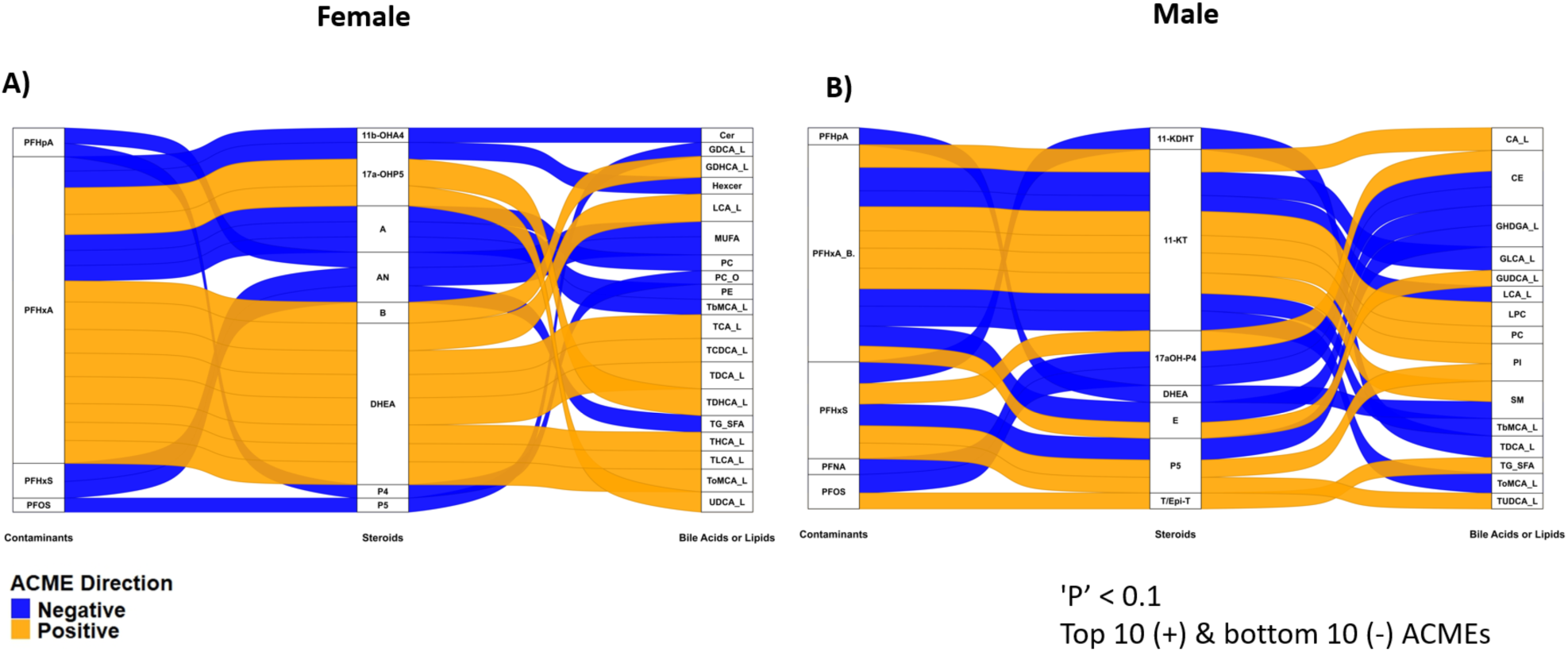
**The causal mediation analysis results showing the indirectly mediated effects with (A) female and (B) male subjects.**

**Fig. S15.**
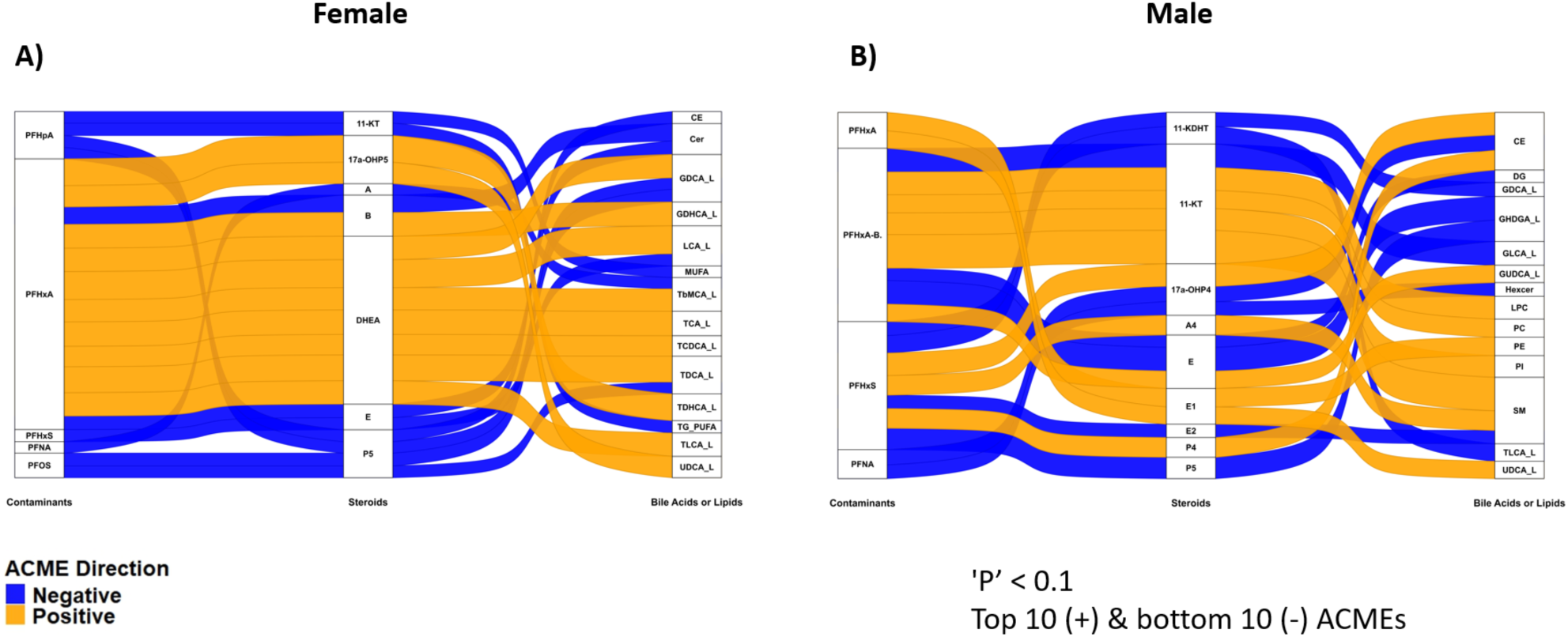
**The causal mediation analysis results showing the indirectly mediated effects with (A) female and (B) male subjects after basic adjustments (age and gender) of the models (Y and M).**

## Supplementary tables

**Table S1.** Demographic characteristics of the study population (median, range), divided into three groups (all, females and males).

| Median (Q1, Q3)<br>(or min and max) | All Subjects | Female Subjects | Male Subjects |
| --- | --- | --- | --- |
| N | 104 | 69 | 35 |
| Age | 49.0 (43.0, 54.3) | 46.0 (43.0, 53.0) | 52.0 (44.5, 58.0) |
| BMI | 45.3 (41.1, 49.1) | 45.11 (41.1, 48.7) | 46.3 (41.3, 49.6) |
| PFAS | 0.41 (0.28, 0.57) | 0.35 (0.25, 0.53) | 0.48 (0.41, 0.70) |
| HDL | 1.1 (1.0, 1.3) | 1.1 (1.0, 1.4) | 1.0 (0.9, 1.2) |
| HOMA-IR | 3.1 (1.8, 4.6) | 3.0 (1.6, 3.9) | 3.9 (2.3, 5.4) |
| Steatosis Grade (min, max) | (0, 3) | (0, 3) | (0, 1) |
| Fibrosis Stage (min, max) | (0, 4) | (0, 3) | (0, 4) |
| Necroinflammation (min, max) | (0, 2) | (0, 1) | (0, 2) |
| HOMA-IR (min, max) | (0.2, 14.5) | (0.2, 14.5) | (0.2, 14.5) |

**Table S2.** The internal standard mix prepared in 30% acetonitrile/70% water with following concentrations.

| Compound | Concentration (nM) |
| --- | --- |
| d3-17a-OH-Pregnenolone (d3-17a-OHP5) | 20 |
| d4-Pregnenolone (d4-P5) | 200 |
| d7-11-Deoxycortisol (d7-S) | 20 |
| d7-11-Deoxycorticosterone (d7-DOC) | 20 |
| d4-Cortisol (d4-F) | 20 |
| d9-Progesterone (d9-P4) | 20 |
| d8-Cortisone (d8-E) | 20 |
| d8-17a-OH-Progesterone (d8-17aOH-P4) | 20 |
| d4-Estradiol (d4-E2) | 20 |
| d4-Estrone (d4-E1) | 20 |
| d4-Androsterone (d4-AN) | 20 |
| d3-Testosterone (d3- T/Epi-T) | 4 |
| d8-Aldosterone (d8-A) | 20 |
| d3-11-Ketotestosterone (d3-11-KT) | 20 |
| d3-11-Ketodihydrotestosterone (d3-11-KDHT) | 20 |
| d4-11b-OH-Androstenedione (d4-11b-OHA4) | 20 |
| d4-Dihydrotestosterone (d4-DHT) | 20 |
| d2-Dehydroepiandrosterone (d2-DHEA) | 200 |
| d3-Androstenedione (d3-A4) | 2 |

**Table S3.** Calibration curve levels.

| Level | Working solution, nM | Reconstituted LC-MS/MS sample (100 μL), pM | Calibration points conc. pM |
| --- | --- | --- | --- |
| U1 | 0.005 | 2 | 1.00 |
| U2 | 0.01 | 4 | 2.00 |
| L1 | 0.025 | 10 | 5.00 |
| L2 | 0.05 | 20 | 10.00 |
| L3 | 0.1 | 40 | 20.00 |
| L4 | 0.25 | 100 | 50.00 |
| L5 | 0.5 | 200 | 100.00 |
| L6 | 1 | 400 | 200.00 |
| L7 | 2.5 | 1000 | 500.00 |
| L8 | 5 | 2000 | 1000.00 |
| L9 | 10 | 4000 | 2000.00 |
| L10 | 50 | 20000 | 10000.00 |
| L11 | 100 | 40000 | 20000.00 |
| L12 | 250 | 100000 | 50000.00 |
| L13 | 500 | 200000 | 100000.00 |
| L14 | 1000 | 400000 | 200000.00 |

**Table S4.**
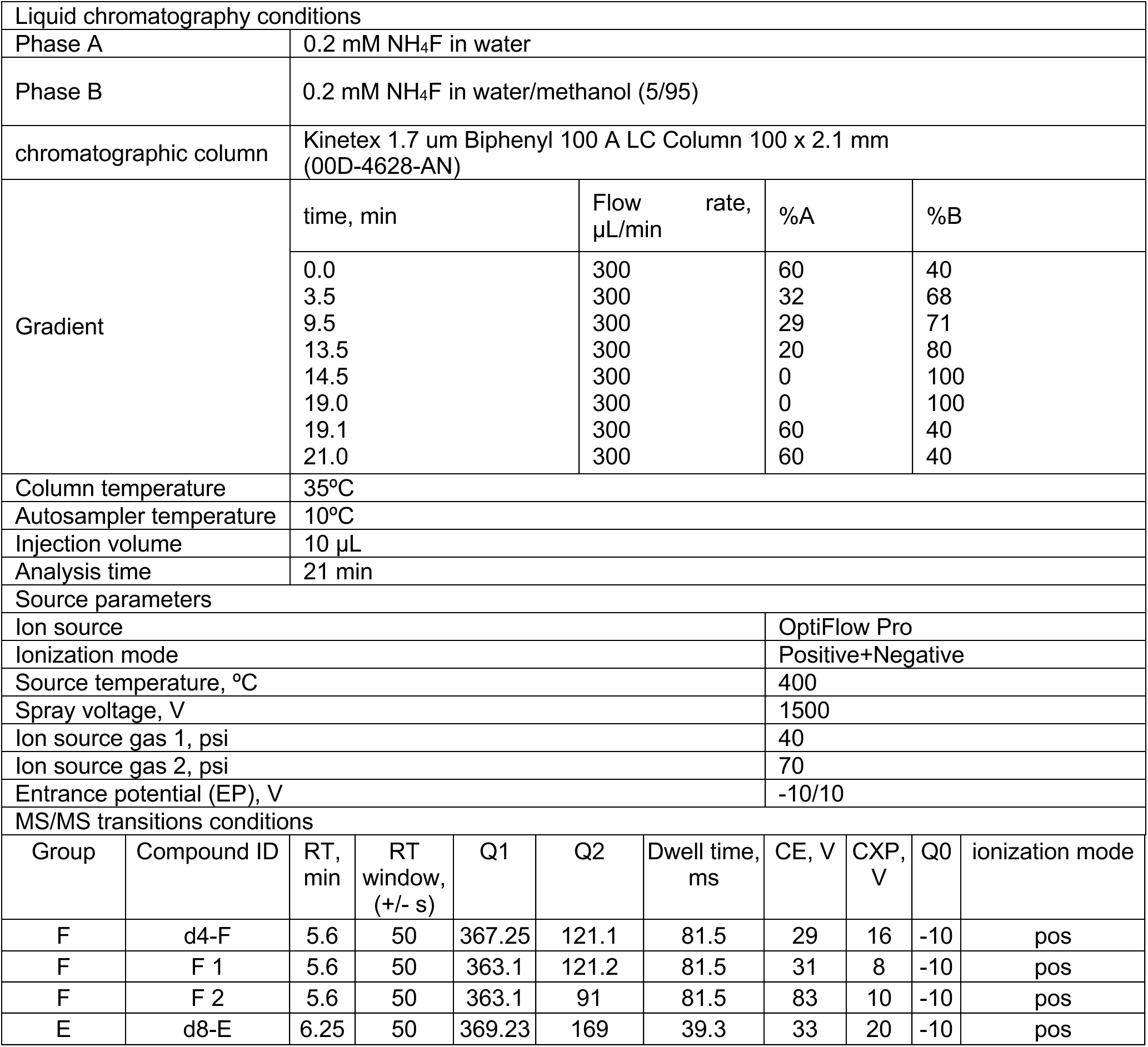
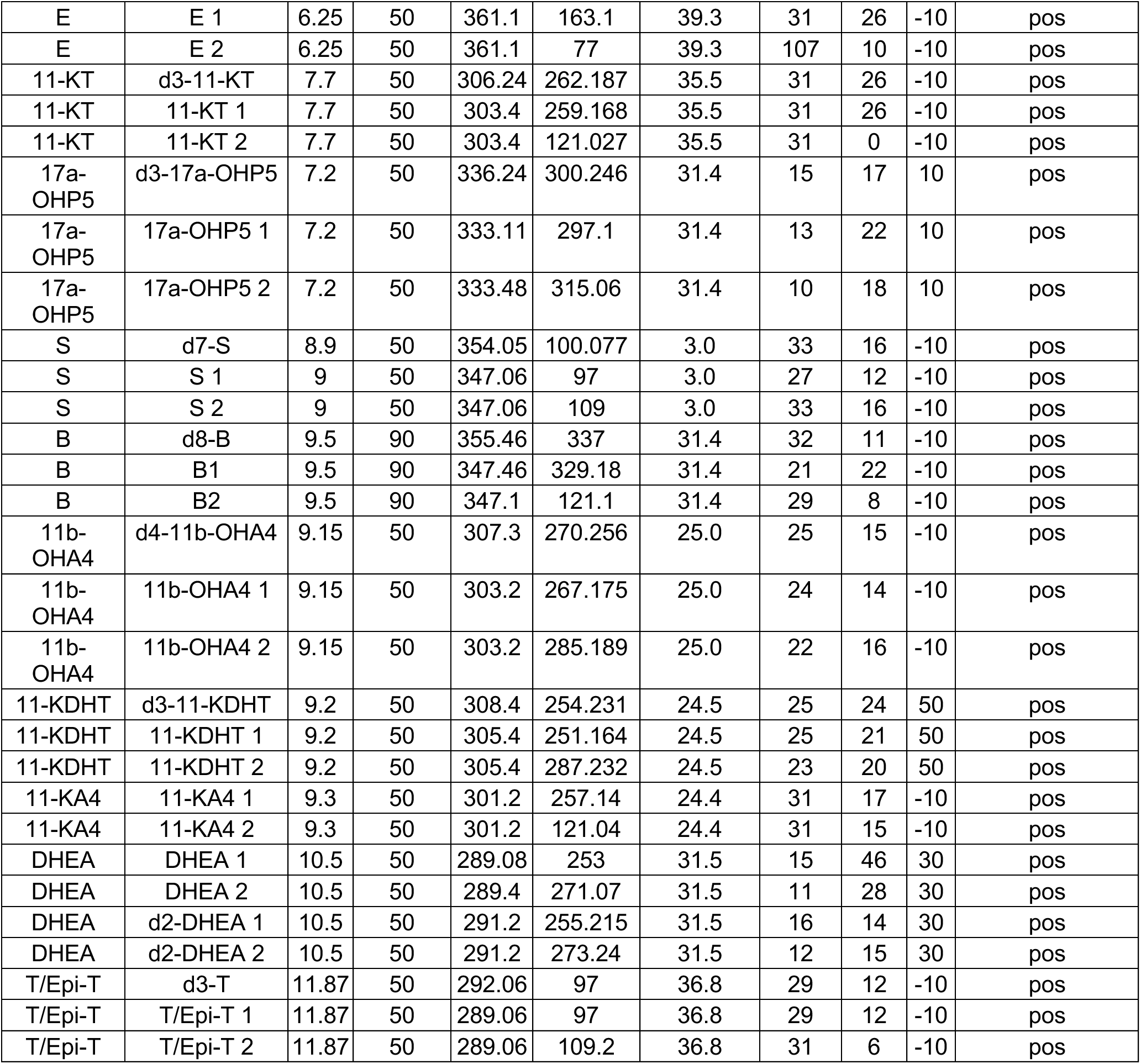
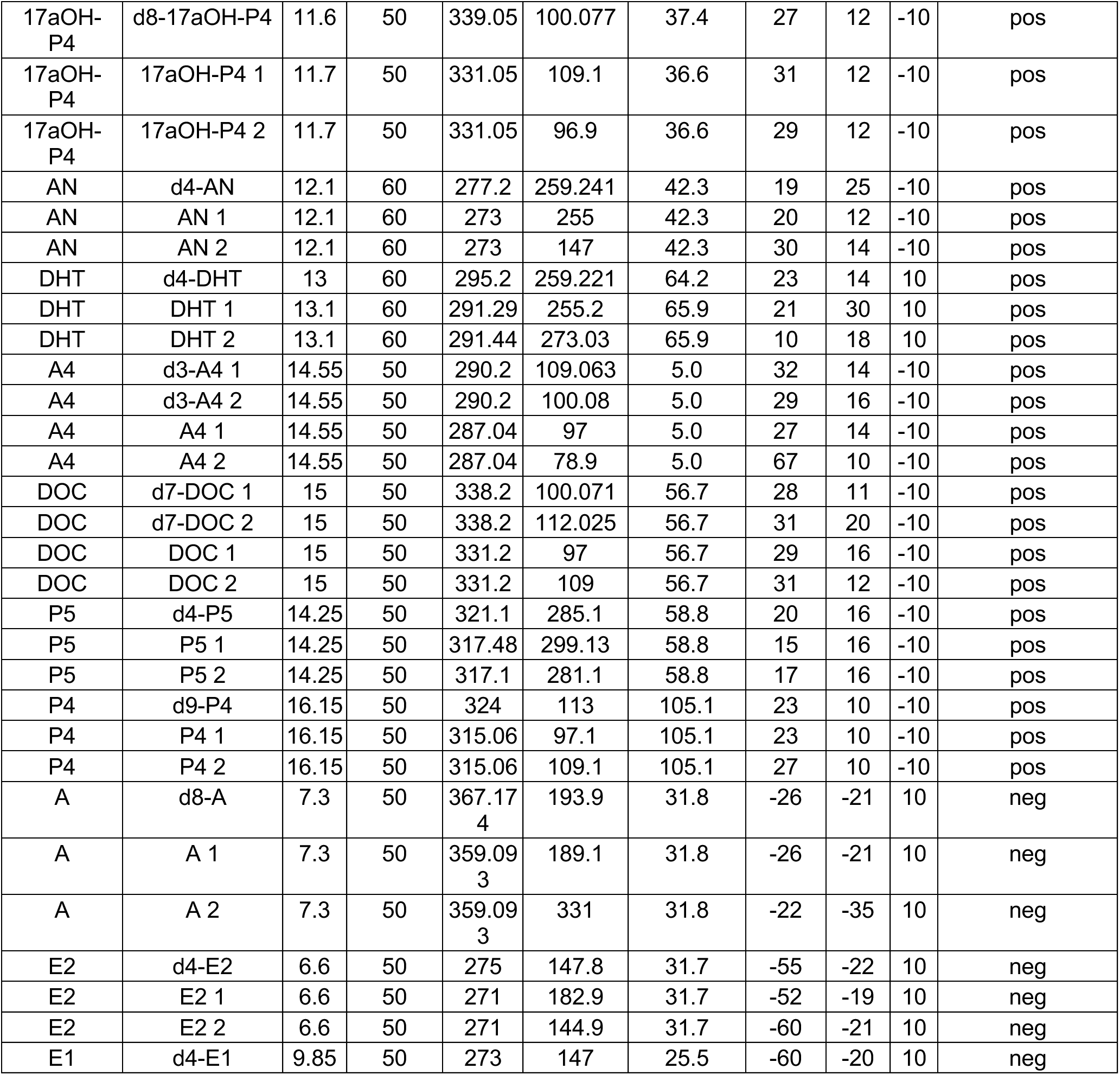
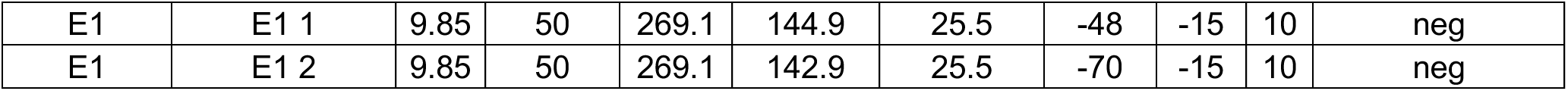
UHPLC-MS/MS conditions for steroid analysis.

**Table S5.** Instrumentation.

| Instrument Model | Serial Number |
| --- | --- |
| ExionLC AD Pump | AB3AD5976127 |
| ExionLC AD Pump | AB3AD5976128 |
| ExionLC AD Autosampler | AB3AC5972305 |
| AC Column Oven | AB2CT5972057 |
| System Controller | ABCBM5974024 |
| SCIEX Triple Quad™ 7500 | FA220602109 |

**Table S6.** The medians of steroid concentrations (pM) with interquartile ranges (Q1 and Q3).

| Steroid Name | Abbreviation of Steroid | Steroid Category | Median (Q1, Q3) (pM) | Median (Q1, Q3) (pM) | Median (Q1, Q3) (pM) |
| --- | --- | --- | --- | --- | --- |
| 11-Ketotestosterone | 11-KT | Androgens | 926 (617, 1161) | 925 (606, 1163) | 942 (759, 1087) |
| 11b-OH-Androstenedione | 11b-OHA4 | Androgens | 8870 (6700, 11868) | 9461 (6379, 11406) | 8663 (7510, 12528) |
| 11-Ketodihydrotestosterone | 11-KDHT | Androgens | 100 (100, 100) | 100 (100, 100) | 100 (100, 153) |
| 11-Ketoandrostenedione | 11-KA4 | Androgens | 1262 (845, 1687) | 1262 (838, 1663) | 1318 (890, 1699) |
| Dehydroepiandrosterone | DHEA | Androgens | 8294 (5602, 12330) | 9395 (6072, 13100) | 6683 (5301, 9524) |
| Testosterone / Epi-testosterone | T/Epi-T | Androgens | 1225 (838, 8089) | 913 (717, 1216) | 10808 (8204, 13367) |
| Androsterone | AN | Androgens | 250 (250, 507) | 250 (250, 505) | 250 (250, 410) |
| Dihydrotestosterone | DHT | Androgens | 276 (132, 461) | 203 (109, 311) | 524 (381, 632) |
| Androstenedione | A4 | Androgens | 2226 (1548, 3064) | 2366 (1578, 3276) | 1995 (1539, 2424) |
| Estradiol | E2 | Estrogens | 137 (77, 271) | 167 (66, 441) | 117 (104, 175) |
| Estrone | E1 | Estrogens | 271 (170, 407) | 321 (193, 482) | 232 (166, 285) |
| Cortisone | E | Glucocorticoids | 40685 (33628, 45755) | 40538 (30857, 45250) | 42424 (37906, 49608) |
| 11-Deoxycortisol | S | Glucocorticoids | 843 (560, 1134) | 810 (533, 1050) | 892 (576, 1346) |
| Corticosterone | B | Mineralocorticoids | 7258 (5046, 12538) | 7545 (5054, 12526) | 6495 (5085, 12852) |
| Aldosterone | A | Mineralocorticoids | 1105 (685, 1827) | 1291 (925, 2004) | 722 (443, 1142) |
| 11-Deoxycorticosterone | DOC | Mineralocorticoids | 62 (47, 90) | 60 (48, 96) | 67 (44, 87) |
| 17a-OH-Pregnenolone | 17a-OHP5 | Progestogens | 1613 (924, 3290) | 1615 (776, 3293) | 1612 (1190, 3050) |
| 17a-OH-Progesterone | 17a-OHP4 | Progestogens | 1121 (631, 1717) | 830 (527, 1510) | 1510 (1076, 1839) |
| Pregnenolone | P5 | Progestogens | 2091 (1288, 4116) | 2375 (1370, 4172) | 1584 (1120, 2464) |
| Progesterone | P4 | Progestogens | 124 (79, 262) | 160 (92, 586) | 94 (75, 146) |

**Table S7.** The medians of PFAS, bile acid and lipid concentrations (pM) with interquartile ranges (Q1 and Q3).

|  | All | Female | Male |
| --- | --- | --- | --- |
| Compounds | Median (Q1, Q3) (pM) | Median (Q1, Q3) (pM) | Median (Q1, Q3) (pM) |
| PFHpA | 0.05 (0.04, 0.08) | 0.05 (0.03, 0.07) | 0.08 (0.06, 0.09) |
| PFHxA | 0.21 (0.18, 0.25) | 0.20 (0.18, 0.25) | 0.23 (0.18, 0.26) |
| PFHxA_Branched | 2.47 (1.55, 5.48) | 2.45 (1.50, 4.59) | 2.77 (1.67, 7.06) |
| PFHxS | 0.04 (0.03, 0.05) | 0.03 (0.03, 0.05) | 0.05 (0.04, 0.07) |
| PFNA | 0.03 (0.02, 0.03) | 0.02 (0.02, 0.03) | 0.03 (0.02, 0.04) |
| PFOA | 0.13 (0.09, 0.16) | 0.13 (0.08, 0.16) | 0.13 (0.10, 0.17) |
| PFOS | 0.14 (0.10, 0.24) | 0.12 (0.09, 0.21) | 0.18 (0.15, 0.27) |
| CA | 11 (7, 16) | 11 (7, 16) | 11 (7, 16) |
| CDCA | 31 (27, 37) | 31 (27, 37) | 30 (28, 43) |
| DCA | 21 (17, 26) | 22 (18, 26) | 19 (16, 26) |
| LCA | 94 (84, 100) | 95 (85, 100) | 90 (84, 102) |
| UDCA | 7 (5, 11) | 9 (6, 13) | 7 (4, 8) |
| GCA | 15967 (9475, 23383) | 17187 (9272, 23102) | 14579 (10409, 22390) |
| GCDCA | 14801 (8961, 23899) | 15554 (8549, 24877) | 12739 (9313, 21418) |
| GDCA | 7438 (3298, 14389) | 7835 (4314, 15531) | 5828 (1410, 12555) |
| GDHCA | 2 (2, 3) | 2 (2, 3) | 2 (2, 3) |
| GHCA | 22 (12, 38) | 25 (14, 40) | 19 (10, 30) |
| GHDGA | 3 (0, 7) | 3 (0, 6) | 3 (0, 8) |
| GLCA | 341 (182, 607) | 354 (214, 590) | 310 (157, 656) |
| GUDCA | 1184 (565, 2334) | 1238 (588, 2503) | 962 (546, 1773) |
| TbMCA | 29 (11, 81) | 33 (13, 80) | 26 (11, 80) |
| ToMCA | 17 (14, 25) | 18 (14, 25) | 17 (12, 24) |
| TCA | 4700 (3054, 6914) | 4387 (2831, 6474) | 6000 (3680, 7981) |
| TCDCA | 4950 (3174, 7237) | 4772 (2966, 7203) | 5351 (3712, 7027) |
| TDCA | 1559 (857, 3996) | 1696 (875, 3897) | 1423 (629, 4361) |
| TDHCA | 11 (10, 14) | 11 (10, 14) | 12 (10, 14) |
| THCA | 18 (15, 24) | 18 (15, 24) | 17 (15, 21) |
| TLCA | 127 (67, 244) | 135 (70, 250) | 124 (63, 240) |
| TUDCA | 116 (73, 214) | 112 (71, 215) | 118 (79, 214) |
| C4 | 35 (19, 66) | 35 (21, 70) | 33 (17, 48) |
| Cer | 33 (28, 38) | 33 (28, 38) | 32 (29, 39) |
| CE | 4 (4, 5) | 4 (4, 5) | 4 (3, 4) |
| DG | 7 (5, 9) | 7 (5, 9) | 7 (6, 9) |
| Hexcer | 7 (6, 8) | 7 (6, 8) | 8 (6, 9) |
| LPC | 6 (5, 7) | 6 (5, 7) | 7 (5, 8) |
| PC | 82 (72, 89) | 83 (72, 89) | 82 (75, 87) |
| PC_O | 6 (5, 7) | 6 (5, 6) | 7 (6, 8) |
| PE | 48 (44, 51) | 46 (43, 50) | 49 (47, 53) |
| PI | 7 (6, 8) | 7 (6, 8) | 7 (6, 8) |
| SM | 20 (18, 23) | 19 (18, 22) | 22 (19, 25) |
| TG_SFA | 53 (36, 82) | 57 (37, 79) | 47 (36, 88) |
| MUFA | 36 (23, 51) | 34 (21, 50) | 43 (30, 57) |
| TG_PUFA | 31 (23, 46) | 28 (21, 40) | 33 (25, 51) |

**Table S8.**
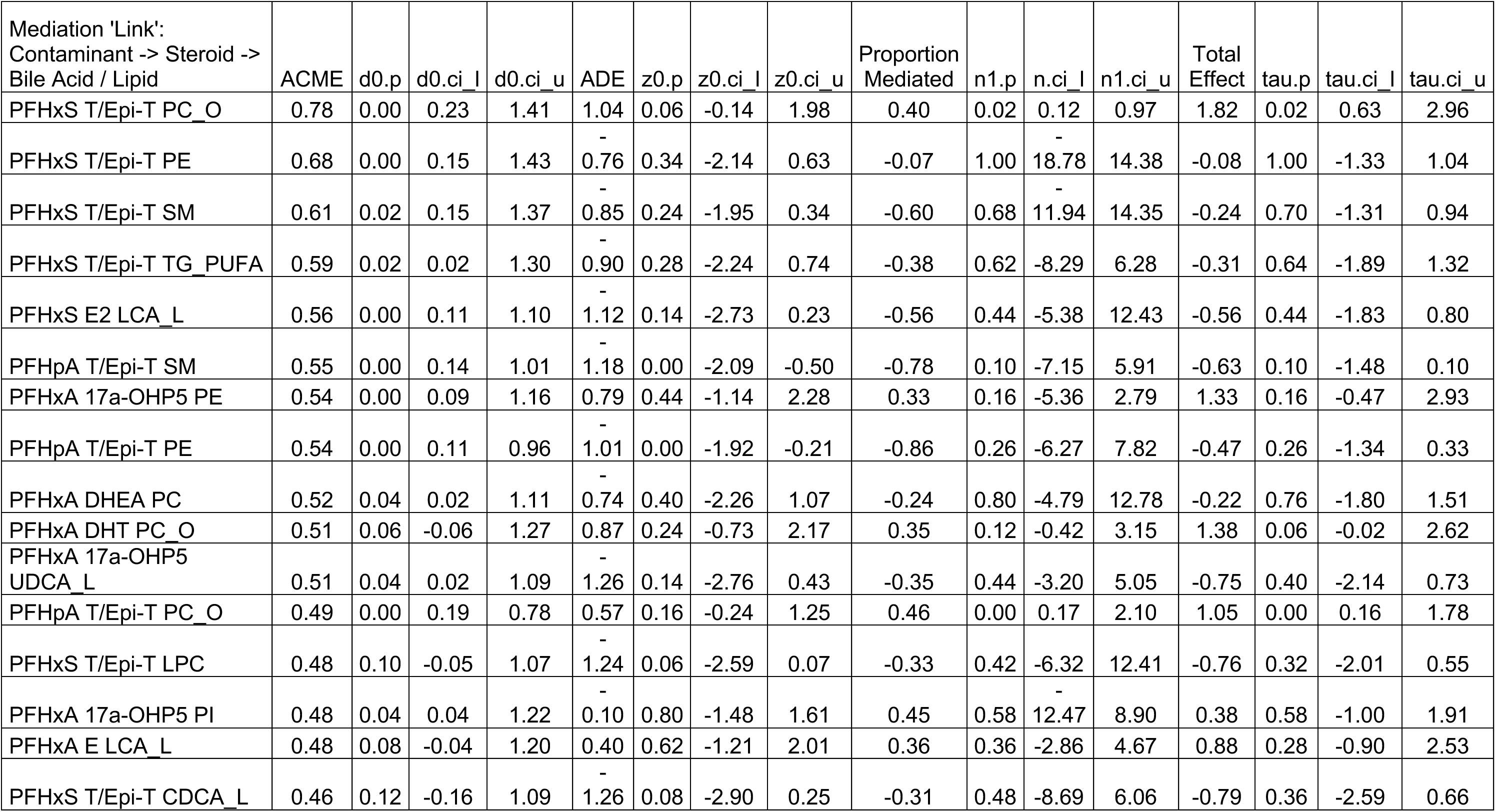
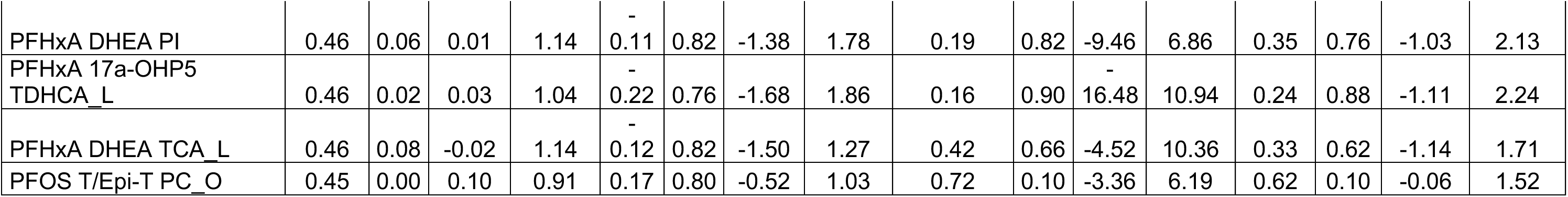
Top 20 unadjusted mediation analysis results as per descending indirect effect (ACME) order of magnitude using all subjects. The calculation for the link was done assuming that mediation happens in the liver. Liver bile acids were also in serum, hence the ‘_L’ at the end to differentiate. Definitions: ADE = average direct effects, the p values, d0.p for ACME and similarly for others (ADE, proportion mediated and total effect), are fraction of simulations that are above or below zero, where the lowest one is selected and multiplied with two, and ci_l=lower confidence interval and ci_u=upper confidence interval.

**Table S9.** Top 20 unadjusted mediation analysis results as per descending indirect effect (ACME) order of magnitude using female subjects. The calculation for the link was done assuming that mediation happens in the liver. Liver bile acids were also in serum, hence the ‘_L’ at the end to differentiate. Definitions: ADE = average direct effects, the p values, d0.p for ACME and similarly for others (ADE, proportion mediated and total effect), are fraction of simulations that are above or below zero, where the lowest one is selected and multiplied with two, and ci_l=lower confidence interval and ci_u=upper confidence interval.

| Mediation 'Link':<br>Contaminant -> Steroid -> Bile<br>Acid / Lipid | ACME | d0.p | d0.ci_l | d0.ci_u | ADE | z0.p | z0.ci_l | z0.ci_u | Proportion<br>Mediated | n1.p | n.ci_l | n1.ci_u | Total<br>Effect | tau.p | tau.ci_l | tau.ci_u |
| --- | --- | --- | --- | --- | --- | --- | --- | --- | --- | --- | --- | --- | --- | --- | --- | --- |
| PFHxA DHEA LCA_L | 0.86 | 0.02 | 0.25 | 1.77 | 0.04 | 0.96 | -1.56 | 1.48 | 0.84 | 0.22 | -5.62 | 19.73 | 0.91 | 0.20 | -0.55 | 2.55 |
| PFHxA DHEA TDCA_L | 0.85 | 0.04 | 0.07 | 1.78 | -0.79 | 0.42 | -2.58 | 1.01 | 0.03 | 1.00 | -20.23 | 6.04 | 0.06 | 0.96 | -1.78 | 1.55 |
| PFHxA 17a-OHP5 TDHCA_L | 0.84 | 0.00 | 0.27 | 1.52 | 0.08 | 0.84 | -1.80 | 1.86 | 0.62 | 0.28 | -4.58 | 7.61 | 0.92 | 0.28 | -1.04 | 2.71 |
| PFHxA DHEA TCA_L | 0.75 | 0.00 | 0.15 | 1.79 | -0.44 | 0.60 | -2.36 | 1.68 | 0.28 | 0.86 | -14.60 | 22.70 | 0.31 | 0.86 | -1.45 | 2.51 |
| PFHxA DHEA TCDCA_L | 0.74 | 0.04 | 0.09 | 1.75 | -0.47 | 0.66 | -2.70 | 1.19 | 0.46 | 0.72 | -7.41 | 8.38 | 0.27 | 0.68 | -1.49 | 1.84 |
| PFHxA DHEA ToMCA_L | 0.72 | 0.08 | -0.06 | 1.47 | -0.29 | 0.78 | -2.20 | 1.56 | 0.45 | 0.66 | -5.35 | 10.35 | 0.43 | 0.62 | -1.39 | 2.22 |
| PFHxA B GDHCA_L | 0.66 | 0.02 | 0.02 | 1.48 | -0.98 | 0.24 | -2.49 | 0.76 | -0.46 | 0.64 | -27.91 | 7.60 | -0.32 | 0.62 | -1.79 | 1.51 |
| PFHxA DHEA GDCA_L | 0.64 | 0.10 | -0.05 | 1.96 | -0.59 | 0.54 | -2.21 | 1.51 | -0.09 | 0.94 | -8.06 | 7.16 | 0.05 | 0.92 | -1.86 | 2.09 |
| PFHxA 17a-OHP5 UDCA_L | 0.63 | 0.04 | 0.01 | 1.54 | -0.47 | 0.50 | -2.05 | 1.25 | 0.17 | 0.90 | -13.09 | 29.91 | 0.17 | 0.90 | -1.49 | 1.52 |
| PFHxA DHEA THCA_L | 0.59 | 0.08 | -0.03 | 1.40 | -0.32 | 0.70 | -2.05 | 1.78 | 0.34 | 0.78 | -4.47 | 6.73 | 0.27 | 0.78 | -1.60 | 2.26 |
| PFHxA DHEA TLCA_L | 0.57 | 0.06 | 0.01 | 1.37 | -1.01 | 0.20 | -2.39 | 0.49 | -0.39 | 0.64 | -13.40 | 8.62 | -0.44 | 0.58 | -1.87 | 1.00 |
| PFHxA E LCA_L | 0.56 | 0.02 | 0.05 | 1.37 | 0.53 | 0.64 | -1.72 | 2.17 | 0.34 | 0.24 | -0.83 | 8.90 | 1.08 | 0.22 | -1.03 | 2.82 |
| PFHxS E2 LCA_L | 0.51 | 0.00 | 0.13 | 1.10 | -0.25 | 0.58 | -1.22 | 0.76 | 0.63 | 0.68 | -5.45 | 7.31 | 0.26 | 0.68 | -0.78 | 1.40 |
| PFHxA E GDHCA_L | 0.50 | 0.14 | -0.06 | 1.44 | -1.01 | 0.40 | -2.92 | 1.18 | -0.18 | 0.66 | -4.29 | 10.63 | -0.51 | 0.68 | -2.34 | 1.40 |
| PFHxA B GLCA_L | 0.50 | 0.08 | 0.00 | 1.32 | 0.69 | 0.52 | -1.12 | 2.36 | 0.29 | 0.24 | -1.38 | 3.34 | 1.19 | 0.16 | -0.53 | 2.95 |
| PFHxA 17a-OHP5 LCA_L | 0.50 | 0.22 | -0.43 | 1.32 | 0.58 | 0.52 | -1.19 | 2.63 | 0.33 | 0.44 | -1.30 | 2.17 | 1.08 | 0.26 | -0.76 | 3.05 |
| PFHxA 11b-OHA4 TG PUFA | 0.48 | 0.06 | -0.01 | 1.14 | -0.53 | 0.56 | -2.27 | 1.16 | -0.09 | 0.88 | -18.04 | 5.37 | -0.05 | 0.90 | -1.85 | 1.59 |
| PFHxA DHEA GLCA_L | 0.48 | 0.16 | -0.15 | 1.35 | 0.57 | 0.52 | -1.22 | 2.30 | 0.29 | 0.44 | -7.12 | 4.57 | 1.04 | 0.28 | -0.59 | 2.73 |
| PFHxA 17a-OHP5 GHDGA_L | 0.46 | 0.16 | -0.08 | 1.32 | -2.48 | 0.00 | -4.13 | -0.81 | -0.21 | 0.16 | -0.96 | 0.04 | -2.03 | 0.00 | -3.64 | -0.60 |
| PFHxA DHEA TbMCA_L | 0.45 | 0.26 | -0.24 | 1.57 | -0.67 | 0.48 | -2.54 | 1.11 | -0.22 | 0.80 | -5.85 | 35.43 | -0.22 | 0.82 | -1.94 | 1.57 |

**Table S10.** Top 20 unadjusted mediation analysis results as per descending indirect effect (ACME) order of magnitude using male subjects. The calculation for the link was done assuming that mediation happens in the liver. Liver bile acids were also in serum, hence the ‘_L’ at the end to differentiate. Definitions: ADE = average direct effects, the p values, d0.p for ACME and similarly for others (ADE, proportion mediated and total effect), are fraction of simulations that are above or below zero, where the lowest one is selected and multiplied with two, and ci_l=lower confidence interval and ci_u=upper confidence interval.

| Mediation 'Link':<br>Contaminant -> Steroid -> Bile<br>Acid / Lipid | ACME | d0.p | d0.ci_<br>l | d0.ci_u | ADE | z0.p | z0.ci_<br>l | z0.ci_u | Proportion<br>Mediated | n1.p | n.ci_l | n1.ci_u | Total<br>Effect | tau.p | tau.ci_<br>l | tau.ci_u |
| --- | --- | --- | --- | --- | --- | --- | --- | --- | --- | --- | --- | --- | --- | --- | --- | --- |
| PFHxA_Branched 11-KT LPC | 0.85 | 0.04 | 0.20 | 1.77 | -1.02 | 0.10 | -2.41 | 0.20 | -0.49 | 0.84 | -14.70 | 11.23 | -0.17 | 0.80 | -1.67 | 1.06 |
| PFHxA_Branched 11-KT CA_L | 0.80 | 0.02 | 0.10 | 1.61 | -1.54 | 0.04 | -3.08 | -0.14 | -0.79 | 0.30 | -22.21 | 17.06 | -0.74 | 0.28 | -2.39 | 0.54 |
| PFHxS 17aOH-P4 CE | 0.73 | 0.02 | 0.03 | 1.86 | 0.60 | 0.58 | -1.11 | 2.60 | 0.48 | 0.22 | -2.20 | 2.88 | 1.33 | 0.20 | -0.52 | 3.19 |
| PFHxA_Branched 11-KT SM | 0.73 | 0.06 | -0.03 | 1.48 | -0.42 | 0.46 | -1.94 | 1.17 | 0.82 | 0.66 | -7.46 | 15.85 | 0.30 | 0.64 | -0.91 | 1.82 |
| PFHxA_Branched 11-KT PI | 0.71 | 0.04 | 0.04 | 1.65 | -0.08 | 0.84 | -1.38 | 1.19 | 0.78 | 0.36 | -5.35 | 12.14 | 0.63 | 0.32 | -0.65 | 1.89 |
| PFHxS 17aOH-P4 SM | 0.67 | 0.10 | -0.09 | 1.73 | -0.75 | 0.36 | -2.23 | 0.88 | -0.21 | 0.88 | -15.97 | 9.12 | -0.08 | 0.94 | -1.49 | 1.66 |
| PFNA 11-KT PI | 0.63 | 0.10 | -0.09 | 1.77 | -0.73 | 0.22 | -1.67 | 0.62 | -0.04 | 0.98 | -28.17 | 16.49 | -0.10 | 0.88 | -1.52 | 1.29 |
| PFHxA_Branched 11-KT PC | 0.62 | 0.04 | 0.04 | 1.47 | 0.07 | 0.90 | -1.31 | 1.47 | 0.50 | 0.36 | -8.02 | 7.91 | 0.69 | 0.36 | -0.58 | 2.06 |
| PFHxS P5 PI | 0.61 | 0.06 | -0.01 | 1.62 | -1.30 | 0.16 | -2.96 | 0.41 | -0.39 | 0.46 | -5.42 | 4.70 | -0.68 | 0.40 | -2.36 | 1.27 |
| PFNA 11-KT LPC | 0.57 | 0.12 | -0.06 | 1.63 | -0.85 | 0.28 | -2.90 | 0.66 | -0.25 | 0.74 | -14.37 | 8.34 | -0.28 | 0.74 | -1.83 | 1.17 |
| PFOA P5 GHDGA_L | 0.57 | 0.14 | -0.26 | 1.36 | 0.12 | 0.90 | -1.60 | 1.62 | 0.39 | 0.42 | -7.34 | 8.57 | 0.69 | 0.44 | -0.98 | 2.39 |
| PFHxS P5 PC | 0.57 | 0.10 | -0.09 | 1.42 | -1.16 | 0.26 | -2.63 | 0.39 | -0.39 | 0.52 | -8.19 | 9.23 | -0.59 | 0.42 | -2.18 | 0.82 |
| PFHxA_Branched E GUDCA_L | 0.57 | 0.04 | 0.05 | 1.41 | -1.17 | 0.10 | -2.37 | 0.20 | -0.61 | 0.32 | -31.34 | 3.99 | -0.60 | 0.28 | -1.77 | 0.43 |
| PFOS T/Epi-T TG_SFA | 0.56 | 0.06 | 0.00 | 1.35 | -1.21 | 0.16 | -2.47 | 0.39 | -0.41 | 0.44 | -7.66 | 4.03 | -0.64 | 0.38 | -1.95 | 1.00 |
| PFHxS P5 TUDCA_L | 0.55 | 0.06 | 0.02 | 1.38 | -0.92 | 0.22 | -2.20 | 0.52 | -0.30 | 0.66 | -11.62 | 8.53 | -0.37 | 0.60 | -1.67 | 1.24 |
| PFNA 11-KT PC | 0.55 | 0.08 | -0.14 | 1.17 | -0.76 | 0.30 | -2.01 | 0.51 | -0.26 | 0.88 | -20.19 | 10.70 | -0.21 | 0.80 | -1.68 | 1.42 |
| PFNA 11-KT SM | 0.54 | 0.08 | -0.03 | 1.30 | -0.92 | 0.18 | -2.46 | 0.37 | -0.35 | 0.70 | -10.15 | 6.61 | -0.38 | 0.62 | -2.33 | 0.86 |
| PFHxS 17aOH-P4 Hexcer | 0.54 | 0.18 | -0.25 | 1.56 | -0.35 | 0.72 | -2.25 | 1.30 | 0.16 | 0.88 | -5.72 | 8.36 | 0.19 | 0.82 | -1.78 | 1.91 |
| PFHxS P5 GUDCA_L | 0.54 | 0.10 | -0.08 | 1.42 | -1.71 | 0.02 | -3.20 | -0.12 | -0.37 | 0.18 | -9.11 | 1.33 | -1.18 | 0.12 | -2.86 | 0.35 |
| PFHxS A4 SM | 0.53 | 0.20 | -0.22 | 1.90 | -0.53 | 0.54 | -2.17 | 1.04 | 0.23 | 0.86 | -5.16 | 6.43 | 0.00 | 0.98 | -1.68 | 2.11 |

**Table S11.**
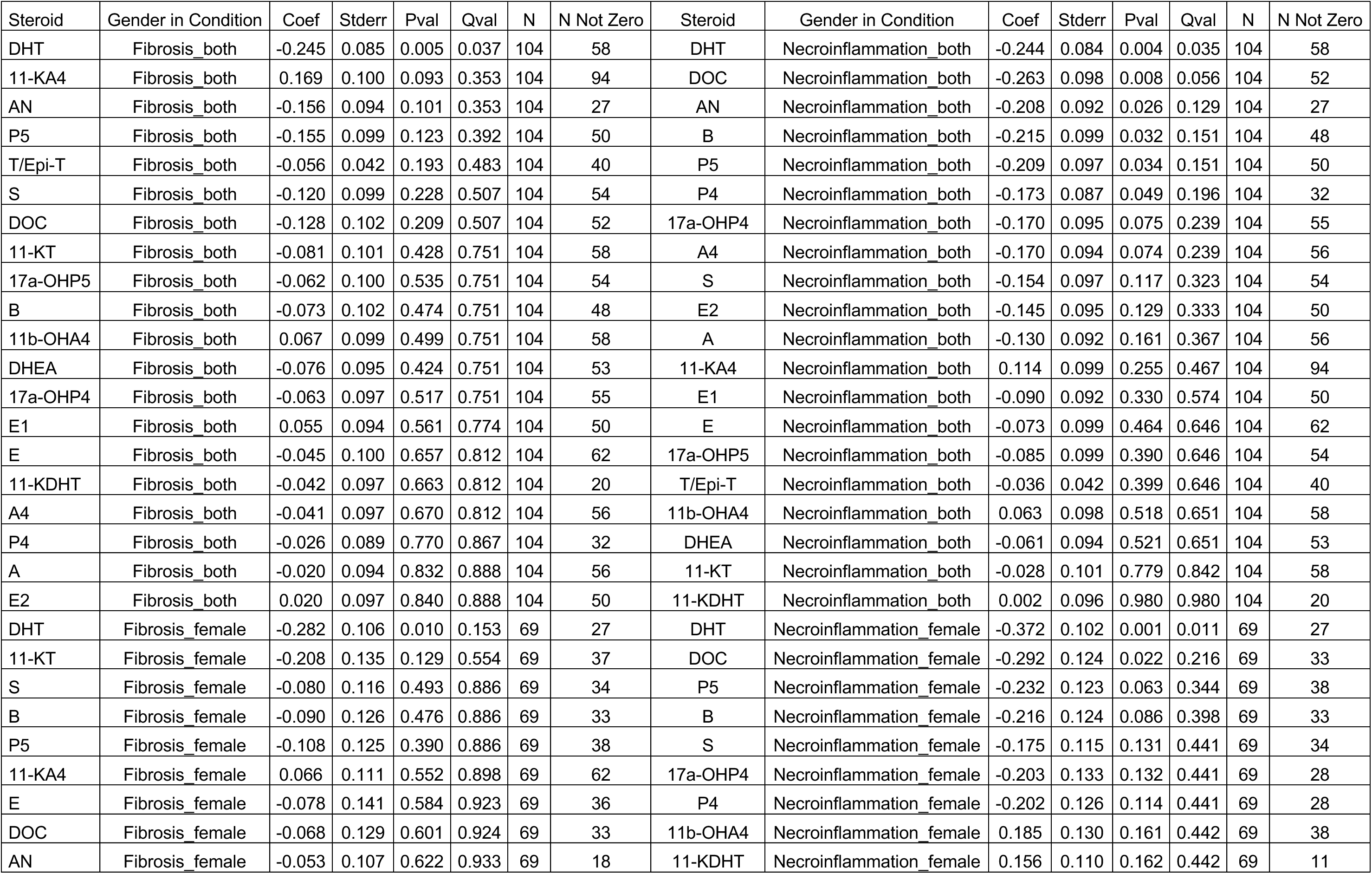
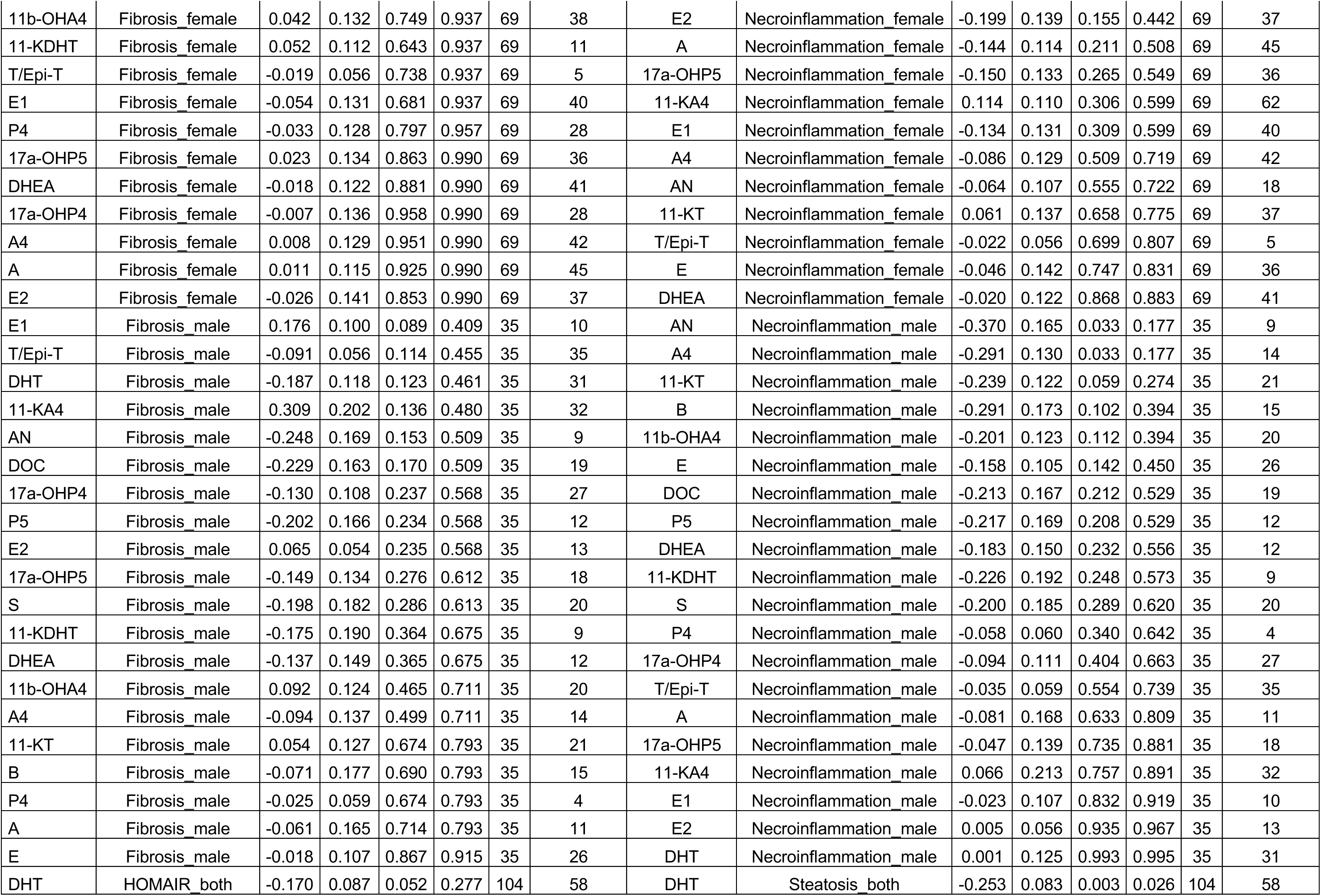
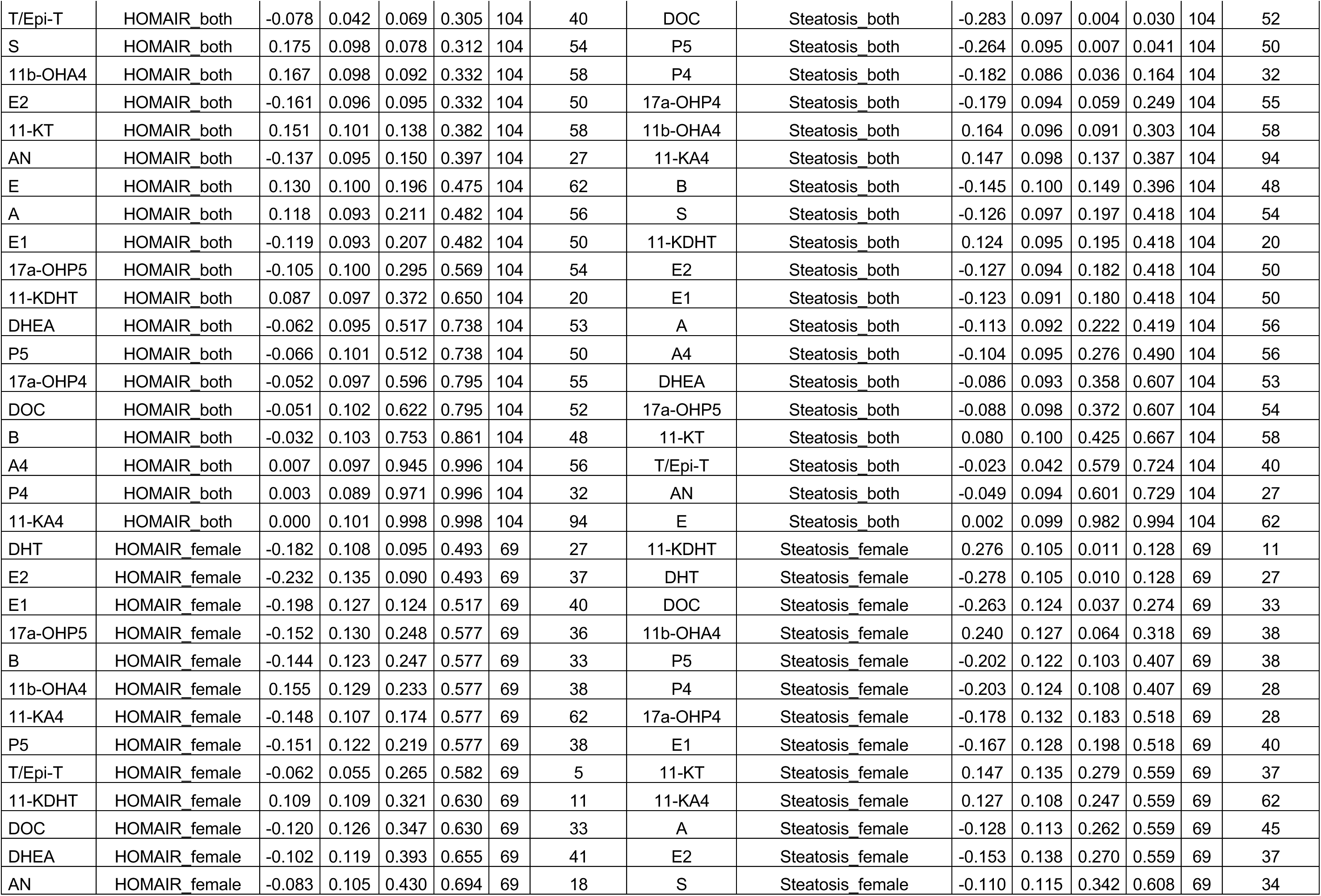
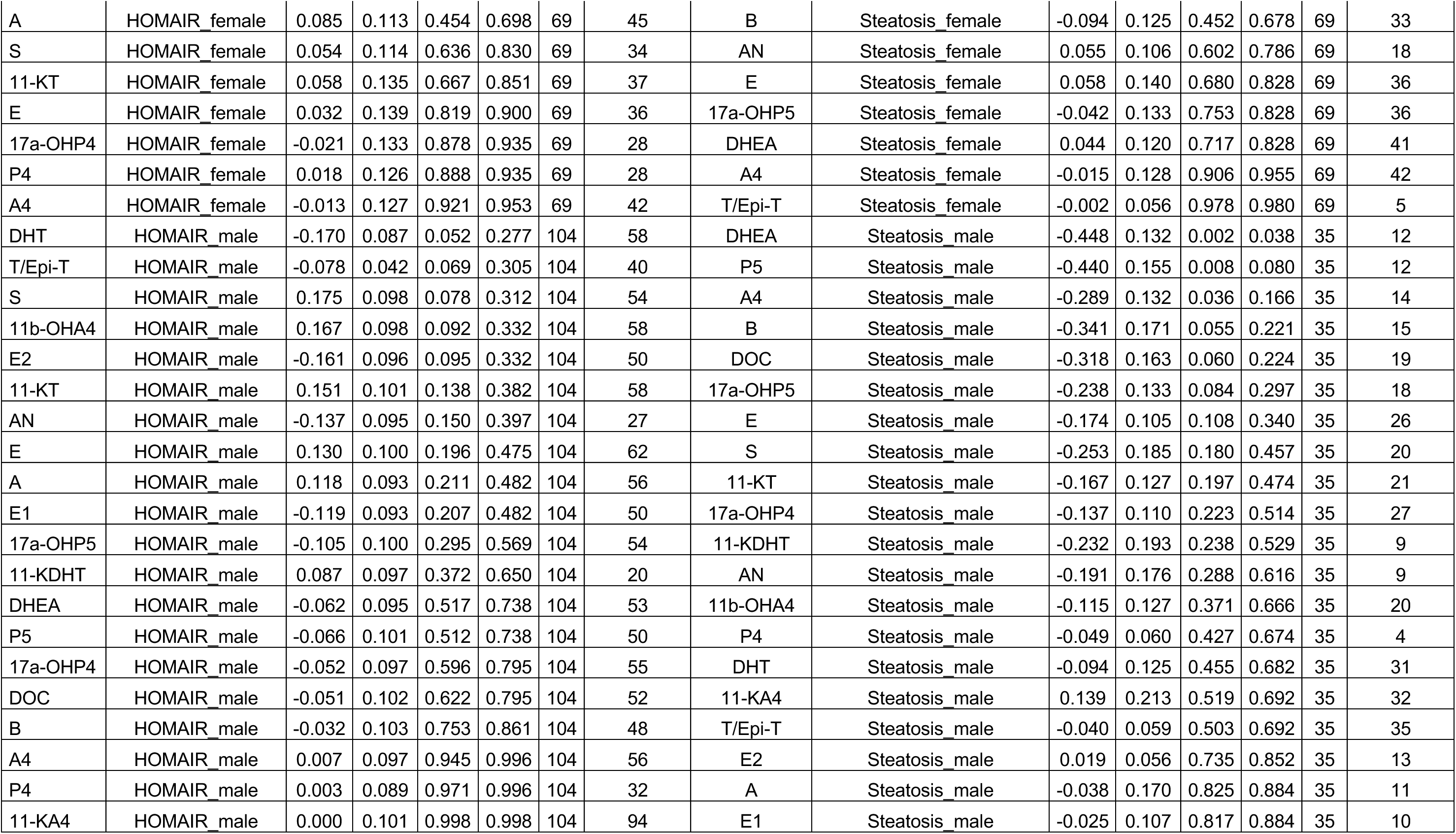
Maaslin-2 linear model results for steroids in cases here in order fibrosis, HOMA-IR, necroinflammation and steatosis.

| Steroid | Gender in Condition | Coef | Stderr | Pval | Qval | N | N Not Zero | Steroid | Gender in Condition | Coef | Stderr | Pval | Qval | N | N Not Zero |
| --- | --- | --- | --- | --- | --- | --- | --- | --- | --- | --- | --- | --- | --- | --- | --- |
| DHT | Fibrosis_both | -0.245 | 0.085 | 0.005 | 0.037 | 104 | 58 | DHT | Necroinflammation_both | -0.244 | 0.084 | 0.004 | 0.035 | 104 | 58 |
| 11-KA4 | Fibrosis_both | 0.169 | 0.100 | 0.093 | 0.353 | 104 | 94 | DOC | Necroinflammation_both | -0.263 | 0.098 | 0.008 | 0.056 | 104 | 52 |
| AN | Fibrosis_both | -0.156 | 0.094 | 0.101 | 0.353 | 104 | 27 | AN | Necroinflammation_both | -0.208 | 0.092 | 0.026 | 0.129 | 104 | 27 |
| P5 | Fibrosis_both | -0.155 | 0.099 | 0.123 | 0.392 | 104 | 50 | B | Necroinflammation_both | -0.215 | 0.099 | 0.032 | 0.151 | 104 | 48 |
| T/Epi-T | Fibrosis_both | -0.056 | 0.042 | 0.193 | 0.483 | 104 | 40 | P5 | Necroinflammation_both | -0.209 | 0.097 | 0.034 | 0.151 | 104 | 50 |
| S | Fibrosis_both | -0.120 | 0.099 | 0.228 | 0.507 | 104 | 54 | P4 | Necroinflammation_both | -0.173 | 0.087 | 0.049 | 0.196 | 104 | 32 |
| DOC | Fibrosis_both | -0.128 | 0.102 | 0.209 | 0.507 | 104 | 52 | 17a-OHP4 | Necroinflammation_both | -0.170 | 0.095 | 0.075 | 0.239 | 104 | 55 |
| 11-KT | Fibrosis_both | -0.081 | 0.101 | 0.428 | 0.751 | 104 | 58 | A4 | Necroinflammation_both | -0.170 | 0.094 | 0.074 | 0.239 | 104 | 56 |
| 17a-OHP5 | Fibrosis_both | -0.062 | 0.100 | 0.535 | 0.751 | 104 | 54 | S | Necroinflammation_both | -0.154 | 0.097 | 0.117 | 0.323 | 104 | 54 |
| B | Fibrosis_both | -0.073 | 0.102 | 0.474 | 0.751 | 104 | 48 | E2 | Necroinflammation_both | -0.145 | 0.095 | 0.129 | 0.333 | 104 | 50 |
| 11b-OHA4 | Fibrosis_both | 0.067 | 0.099 | 0.499 | 0.751 | 104 | 58 | A | Necroinflammation_both | -0.130 | 0.092 | 0.161 | 0.367 | 104 | 56 |
| DHEA | Fibrosis_both | -0.076 | 0.095 | 0.424 | 0.751 | 104 | 53 | 11-KA4 | Necroinflammation_both | 0.114 | 0.099 | 0.255 | 0.467 | 104 | 94 |
| 17a-OHP4 | Fibrosis_both | -0.063 | 0.097 | 0.517 | 0.751 | 104 | 55 | E1 | Necroinflammation_both | -0.090 | 0.092 | 0.330 | 0.574 | 104 | 50 |
| E1 | Fibrosis_both | 0.055 | 0.094 | 0.561 | 0.774 | 104 | 50 | E | Necroinflammation_both | -0.073 | 0.099 | 0.464 | 0.646 | 104 | 62 |
| E | Fibrosis_both | -0.045 | 0.100 | 0.657 | 0.812 | 104 | 62 | 17a-OHP5 | Necroinflammation_both | -0.085 | 0.099 | 0.390 | 0.646 | 104 | 54 |
| 11-KDHT | Fibrosis_both | -0.042 | 0.097 | 0.663 | 0.812 | 104 | 20 | T/Epi-T | Necroinflammation_both | -0.036 | 0.042 | 0.399 | 0.646 | 104 | 40 |
| A4 | Fibrosis_both | -0.041 | 0.097 | 0.670 | 0.812 | 104 | 56 | 11b-OHA4 | Necroinflammation_both | 0.063 | 0.098 | 0.518 | 0.651 | 104 | 58 |
| P4 | Fibrosis_both | -0.026 | 0.089 | 0.770 | 0.867 | 104 | 32 | DHEA | Necroinflammation_both | -0.061 | 0.094 | 0.521 | 0.651 | 104 | 53 |
| A | Fibrosis_both | -0.020 | 0.094 | 0.832 | 0.888 | 104 | 56 | 11-KT | Necroinflammation_both | -0.028 | 0.101 | 0.779 | 0.842 | 104 | 58 |
| E2 | Fibrosis_both | 0.020 | 0.097 | 0.840 | 0.888 | 104 | 50 | 11-KDHT | Necroinflammation_both | 0.002 | 0.096 | 0.980 | 0.980 | 104 | 20 |
| DHT | Fibrosis_female | -0.282 | 0.106 | 0.010 | 0.153 | 69 | 27 | DHT | Necroinflammation_female | -0.372 | 0.102 | 0.001 | 0.011 | 69 | 27 |
| 11-KT | Fibrosis_female | -0.208 | 0.135 | 0.129 | 0.554 | 69 | 37 | DOC | Necroinflammation_female | -0.292 | 0.124 | 0.022 | 0.216 | 69 | 33 |
| S | Fibrosis_female | -0.080 | 0.116 | 0.493 | 0.886 | 69 | 34 | P5 | Necroinflammation_female | -0.232 | 0.123 | 0.063 | 0.344 | 69 | 38 |
| B | Fibrosis_female | -0.090 | 0.126 | 0.476 | 0.886 | 69 | 33 | B | Necroinflammation_female | -0.216 | 0.124 | 0.086 | 0.398 | 69 | 33 |
| P5 | Fibrosis_female | -0.108 | 0.125 | 0.390 | 0.886 | 69 | 38 | S | Necroinflammation_female | -0.175 | 0.115 | 0.131 | 0.441 | 69 | 34 |
| 11-KA4 | Fibrosis_female | 0.066 | 0.111 | 0.552 | 0.898 | 69 | 62 | 17a-OHP4 | Necroinflammation_female | -0.203 | 0.133 | 0.132 | 0.441 | 69 | 28 |
| E | Fibrosis_female | -0.078 | 0.141 | 0.584 | 0.923 | 69 | 36 | P4 | Necroinflammation_female | -0.202 | 0.126 | 0.114 | 0.441 | 69 | 28 |
| DOC | Fibrosis_female | -0.068 | 0.129 | 0.601 | 0.924 | 69 | 33 | 11b-OHA4 | Necroinflammation_female | 0.185 | 0.130 | 0.161 | 0.442 | 69 | 38 |
| AN | Fibrosis_female | -0.053 | 0.107 | 0.622 | 0.933 | 69 | 18 | 11-KDHT | Necroinflammation_female | 0.156 | 0.110 | 0.162 | 0.442 | 69 | 11 |
| 11b-OHA4 | Fibrosis_female | 0.042 | 0.132 | 0.749 | 0.937 | 69 | 38 | E2 | Necroinflammation_female | -0.199 | 0.139 | 0.155 | 0.442 | 69 | 37 |
| 11-KDHT | Fibrosis_female | 0.052 | 0.112 | 0.643 | 0.937 | 69 | 11 | A | Necroinflammation_female | -0.144 | 0.114 | 0.211 | 0.508 | 69 | 45 |
| T/Epi-T | Fibrosis_female | -0.019 | 0.056 | 0.738 | 0.937 | 69 | 5 | 17a-OHP5 | Necroinflammation_female | -0.150 | 0.133 | 0.265 | 0.549 | 69 | 36 |
| E1 | Fibrosis_female | -0.054 | 0.131 | 0.681 | 0.937 | 69 | 40 | 11-KA4 | Necroinflammation_female | 0.114 | 0.110 | 0.306 | 0.599 | 69 | 62 |
| P4 | Fibrosis_female | -0.033 | 0.128 | 0.797 | 0.957 | 69 | 28 | E1 | Necroinflammation_female | -0.134 | 0.131 | 0.309 | 0.599 | 69 | 40 |
| 17a-OHP5 | Fibrosis_female | 0.023 | 0.134 | 0.863 | 0.990 | 69 | 36 | A4 | Necroinflammation_female | -0.086 | 0.129 | 0.509 | 0.719 | 69 | 42 |
| DHEA | Fibrosis_female | -0.018 | 0.122 | 0.881 | 0.990 | 69 | 41 | AN | Necroinflammation_female | -0.064 | 0.107 | 0.555 | 0.722 | 69 | 18 |
| 17a-OHP4 | Fibrosis_female | -0.007 | 0.136 | 0.958 | 0.990 | 69 | 28 | 11-KT | Necroinflammation_female | 0.061 | 0.137 | 0.658 | 0.775 | 69 | 37 |
| A4 | Fibrosis_female | 0.008 | 0.129 | 0.951 | 0.990 | 69 | 42 | T/Epi-T | Necroinflammation_female | -0.022 | 0.056 | 0.699 | 0.807 | 69 | 5 |
| A | Fibrosis_female | 0.011 | 0.115 | 0.925 | 0.990 | 69 | 45 | E | Necroinflammation_female | -0.046 | 0.142 | 0.747 | 0.831 | 69 | 36 |
| E2 | Fibrosis_female | -0.026 | 0.141 | 0.853 | 0.990 | 69 | 37 | DHEA | Necroinflammation_female | -0.020 | 0.122 | 0.868 | 0.883 | 69 | 41 |
| E1 | Fibrosis_male | 0.176 | 0.100 | 0.089 | 0.409 | 35 | 10 | AN | Necroinflammation_male | -0.370 | 0.165 | 0.033 | 0.177 | 35 | 9 |
| T/Epi-T | Fibrosis_male | -0.091 | 0.056 | 0.114 | 0.455 | 35 | 35 | A4 | Necroinflammation_male | -0.291 | 0.130 | 0.033 | 0.177 | 35 | 14 |
| DHT | Fibrosis_male | -0.187 | 0.118 | 0.123 | 0.461 | 35 | 31 | 11-KT | Necroinflammation_male | -0.239 | 0.122 | 0.059 | 0.274 | 35 | 21 |
| 11-KA4 | Fibrosis_male | 0.309 | 0.202 | 0.136 | 0.480 | 35 | 32 | B | Necroinflammation_male | -0.291 | 0.173 | 0.102 | 0.394 | 35 | 15 |
| AN | Fibrosis_male | -0.248 | 0.169 | 0.153 | 0.509 | 35 | 9 | 11b-OHA4 | Necroinflammation_male | -0.201 | 0.123 | 0.112 | 0.394 | 35 | 20 |
| DOC | Fibrosis_male | -0.229 | 0.163 | 0.170 | 0.509 | 35 | 19 | E | Necroinflammation_male | -0.158 | 0.105 | 0.142 | 0.450 | 35 | 26 |
| 17a-OHP4 | Fibrosis_male | -0.130 | 0.108 | 0.237 | 0.568 | 35 | 27 | DOC | Necroinflammation_male | -0.213 | 0.167 | 0.212 | 0.529 | 35 | 19 |
| P5 | Fibrosis_male | -0.202 | 0.166 | 0.234 | 0.568 | 35 | 12 | P5 | Necroinflammation_male | -0.217 | 0.169 | 0.208 | 0.529 | 35 | 12 |
| E2 | Fibrosis_male | 0.065 | 0.054 | 0.235 | 0.568 | 35 | 13 | DHEA | Necroinflammation_male | -0.183 | 0.150 | 0.232 | 0.556 | 35 | 12 |
| 17a-OHP5 | Fibrosis_male | -0.149 | 0.134 | 0.276 | 0.612 | 35 | 18 | 11-KDHT | Necroinflammation_male | -0.226 | 0.192 | 0.248 | 0.573 | 35 | 9 |
| S | Fibrosis_male | -0.198 | 0.182 | 0.286 | 0.613 | 35 | 20 | S | Necroinflammation_male | -0.200 | 0.185 | 0.289 | 0.620 | 35 | 20 |
| 11-KDHT | Fibrosis_male | -0.175 | 0.190 | 0.364 | 0.675 | 35 | 9 | P4 | Necroinflammation_male | -0.058 | 0.060 | 0.340 | 0.642 | 35 | 4 |
| DHEA | Fibrosis_male | -0.137 | 0.149 | 0.365 | 0.675 | 35 | 12 | 17a-OHP4 | Necroinflammation_male | -0.094 | 0.111 | 0.404 | 0.663 | 35 | 27 |
| 11b-OHA4 | Fibrosis_male | 0.092 | 0.124 | 0.465 | 0.711 | 35 | 20 | T/Epi-T | Necroinflammation_male | -0.035 | 0.059 | 0.554 | 0.739 | 35 | 35 |
| A4 | Fibrosis_male | -0.094 | 0.137 | 0.499 | 0.711 | 35 | 14 | A | Necroinflammation_male | -0.081 | 0.168 | 0.633 | 0.809 | 35 | 11 |
| 11-KT | Fibrosis_male | 0.054 | 0.127 | 0.674 | 0.793 | 35 | 21 | 17a-OHP5 | Necroinflammation_male | -0.047 | 0.139 | 0.735 | 0.881 | 35 | 18 |
| B | Fibrosis_male | -0.071 | 0.177 | 0.690 | 0.793 | 35 | 15 | 11-KA4 | Necroinflammation_male | 0.066 | 0.213 | 0.757 | 0.891 | 35 | 32 |
| P4 | Fibrosis_male | -0.025 | 0.059 | 0.674 | 0.793 | 35 | 4 | E1 | Necroinflammation_male | -0.023 | 0.107 | 0.832 | 0.919 | 35 | 10 |
| A | Fibrosis_male | -0.061 | 0.165 | 0.714 | 0.793 | 35 | 11 | E2 | Necroinflammation_male | 0.005 | 0.056 | 0.935 | 0.967 | 35 | 13 |
| E | Fibrosis_male | -0.018 | 0.107 | 0.867 | 0.915 | 35 | 26 | DHT | Necroinflammation_male | 0.001 | 0.125 | 0.993 | 0.995 | 35 | 31 |
| DHT | HOMAIR_both | -0.170 | 0.087 | 0.052 | 0.277 | 104 | 58 | DHT | Steatosis_both | -0.253 | 0.083 | 0.003 | 0.026 | 104 | 58 |
| T/Epi-T | HOMAIR_both | -0.078 | 0.042 | 0.069 | 0.305 | 104 | 40 | DOC | Steatosis_both | -0.283 | 0.097 | 0.004 | 0.030 | 104 | 52 |
| S | HOMAIR_both | 0.175 | 0.098 | 0.078 | 0.312 | 104 | 54 | P5 | Steatosis_both | -0.264 | 0.095 | 0.007 | 0.041 | 104 | 50 |
| 11b-OHA4 | HOMAIR_both | 0.167 | 0.098 | 0.092 | 0.332 | 104 | 58 | P4 | Steatosis_both | -0.182 | 0.086 | 0.036 | 0.164 | 104 | 32 |
| E2 | HOMAIR_both | -0.161 | 0.096 | 0.095 | 0.332 | 104 | 50 | 17a-OHP4 | Steatosis_both | -0.179 | 0.094 | 0.059 | 0.249 | 104 | 55 |
| 11-KT | HOMAIR_both | 0.151 | 0.101 | 0.138 | 0.382 | 104 | 58 | 11b-OHA4 | Steatosis_both | 0.164 | 0.096 | 0.091 | 0.303 | 104 | 58 |
| AN | HOMAIR_both | -0.137 | 0.095 | 0.150 | 0.397 | 104 | 27 | 11-KA4 | Steatosis_both | 0.147 | 0.098 | 0.137 | 0.387 | 104 | 94 |
| E | HOMAIR_both | 0.130 | 0.100 | 0.196 | 0.475 | 104 | 62 | B | Steatosis_both | -0.145 | 0.100 | 0.149 | 0.396 | 104 | 48 |
| A | HOMAIR_both | 0.118 | 0.093 | 0.211 | 0.482 | 104 | 56 | S | Steatosis_both | -0.126 | 0.097 | 0.197 | 0.418 | 104 | 54 |
| E1 | HOMAIR_both | -0.119 | 0.093 | 0.207 | 0.482 | 104 | 50 | 11-KDHT | Steatosis_both | 0.124 | 0.095 | 0.195 | 0.418 | 104 | 20 |
| 17a-OHP5 | HOMAIR_both | -0.105 | 0.100 | 0.295 | 0.569 | 104 | 54 | E2 | Steatosis_both | -0.127 | 0.094 | 0.182 | 0.418 | 104 | 50 |
| 11-KDHT | HOMAIR_both | 0.087 | 0.097 | 0.372 | 0.650 | 104 | 20 | E1 | Steatosis_both | -0.123 | 0.091 | 0.180 | 0.418 | 104 | 50 |
| DHEA | HOMAIR_both | -0.062 | 0.095 | 0.517 | 0.738 | 104 | 53 | A | Steatosis_both | -0.113 | 0.092 | 0.222 | 0.419 | 104 | 56 |
| P5 | HOMAIR_both | -0.066 | 0.101 | 0.512 | 0.738 | 104 | 50 | A4 | Steatosis_both | -0.104 | 0.095 | 0.276 | 0.490 | 104 | 56 |
| 17a-OHP4 | HOMAIR_both | -0.052 | 0.097 | 0.596 | 0.795 | 104 | 55 | DHEA | Steatosis_both | -0.086 | 0.093 | 0.358 | 0.607 | 104 | 53 |
| DOC | HOMAIR_both | -0.051 | 0.102 | 0.622 | 0.795 | 104 | 52 | 17a-OHP5 | Steatosis_both | -0.088 | 0.098 | 0.372 | 0.607 | 104 | 54 |
| B | HOMAIR_both | -0.032 | 0.103 | 0.753 | 0.861 | 104 | 48 | 11-KT | Steatosis_both | 0.080 | 0.100 | 0.425 | 0.667 | 104 | 58 |
| A4 | HOMAIR_both | 0.007 | 0.097 | 0.945 | 0.996 | 104 | 56 | T/Epi-T | Steatosis_both | -0.023 | 0.042 | 0.579 | 0.724 | 104 | 40 |
| P4 | HOMAIR_both | 0.003 | 0.089 | 0.971 | 0.996 | 104 | 32 | AN | Steatosis_both | -0.049 | 0.094 | 0.601 | 0.729 | 104 | 27 |
| 11-KA4 | HOMAIR_both | 0.000 | 0.101 | 0.998 | 0.998 | 104 | 94 | E | Steatosis_both | 0.002 | 0.099 | 0.982 | 0.994 | 104 | 62 |
| DHT | HOMAIR_female | -0.182 | 0.108 | 0.095 | 0.493 | 69 | 27 | 11-KDHT | Steatosis_female | 0.276 | 0.105 | 0.011 | 0.128 | 69 | 11 |
| E2 | HOMAIR_female | -0.232 | 0.135 | 0.090 | 0.493 | 69 | 37 | DHT | Steatosis_female | -0.278 | 0.105 | 0.010 | 0.128 | 69 | 27 |
| E1 | HOMAIR_female | -0.198 | 0.127 | 0.124 | 0.517 | 69 | 40 | DOC | Steatosis_female | -0.263 | 0.124 | 0.037 | 0.274 | 69 | 33 |
| 17a-OHP5 | HOMAIR_female | -0.152 | 0.130 | 0.248 | 0.577 | 69 | 36 | 11b-OHA4 | Steatosis_female | 0.240 | 0.127 | 0.064 | 0.318 | 69 | 38 |
| B | HOMAIR_female | -0.144 | 0.123 | 0.247 | 0.577 | 69 | 33 | P5 | Steatosis_female | -0.202 | 0.122 | 0.103 | 0.407 | 69 | 38 |
| 11b-OHA4 | HOMAIR_female | 0.155 | 0.129 | 0.233 | 0.577 | 69 | 38 | P4 | Steatosis_female | -0.203 | 0.124 | 0.108 | 0.407 | 69 | 28 |
| 11-KA4 | HOMAIR_female | -0.148 | 0.107 | 0.174 | 0.577 | 69 | 62 | 17a-OHP4 | Steatosis_female | -0.178 | 0.132 | 0.183 | 0.518 | 69 | 28 |
| P5 | HOMAIR_female | -0.151 | 0.122 | 0.219 | 0.577 | 69 | 38 | E1 | Steatosis_female | -0.167 | 0.128 | 0.198 | 0.518 | 69 | 40 |
| T/Epi-T | HOMAIR_female | -0.062 | 0.055 | 0.265 | 0.582 | 69 | 5 | 11-KT | Steatosis_female | 0.147 | 0.135 | 0.279 | 0.559 | 69 | 37 |
| 11-KDHT | HOMAIR_female | 0.109 | 0.109 | 0.321 | 0.630 | 69 | 11 | 11-KA4 | Steatosis_female | 0.127 | 0.108 | 0.247 | 0.559 | 69 | 62 |
| DOC | HOMAIR_female | -0.120 | 0.126 | 0.347 | 0.630 | 69 | 33 | A | Steatosis_female | -0.128 | 0.113 | 0.262 | 0.559 | 69 | 45 |
| DHEA | HOMAIR_female | -0.102 | 0.119 | 0.393 | 0.655 | 69 | 41 | E2 | Steatosis_female | -0.153 | 0.138 | 0.270 | 0.559 | 69 | 37 |
| AN | HOMAIR_female | -0.083 | 0.105 | 0.430 | 0.694 | 69 | 18 | S | Steatosis_female | -0.110 | 0.115 | 0.342 | 0.608 | 69 | 34 |
| A | HOMAIR_female | 0.085 | 0.113 | 0.454 | 0.698 | 69 | 45 | B | Steatosis_female | -0.094 | 0.125 | 0.452 | 0.678 | 69 | 33 |
| S | HOMAIR_female | 0.054 | 0.114 | 0.636 | 0.830 | 69 | 34 | AN | Steatosis_female | 0.055 | 0.106 | 0.602 | 0.786 | 69 | 18 |
| 11-KT | HOMAIR_female | 0.058 | 0.135 | 0.667 | 0.851 | 69 | 37 | E | Steatosis_female | 0.058 | 0.140 | 0.680 | 0.828 | 69 | 36 |
| E | HOMAIR_female | 0.032 | 0.139 | 0.819 | 0.900 | 69 | 36 | 17a-OHP5 | Steatosis_female | -0.042 | 0.133 | 0.753 | 0.828 | 69 | 36 |
| 17a-OHP4 | HOMAIR_female | -0.021 | 0.133 | 0.878 | 0.935 | 69 | 28 | DHEA | Steatosis_female | 0.044 | 0.120 | 0.717 | 0.828 | 69 | 41 |
| P4 | HOMAIR_female | 0.018 | 0.126 | 0.888 | 0.935 | 69 | 28 | A4 | Steatosis_female | -0.015 | 0.128 | 0.906 | 0.955 | 69 | 42 |
| A4 | HOMAIR_female | -0.013 | 0.127 | 0.921 | 0.953 | 69 | 42 | T/Epi-T | Steatosis_female | -0.002 | 0.056 | 0.978 | 0.980 | 69 | 5 |
| DHT | HOMAIR_male | -0.170 | 0.087 | 0.052 | 0.277 | 104 | 58 | DHEA | Steatosis_male | -0.448 | 0.132 | 0.002 | 0.038 | 35 | 12 |
| T/Epi-T | HOMAIR_male | -0.078 | 0.042 | 0.069 | 0.305 | 104 | 40 | P5 | Steatosis_male | -0.440 | 0.155 | 0.008 | 0.080 | 35 | 12 |
| S | HOMAIR_male | 0.175 | 0.098 | 0.078 | 0.312 | 104 | 54 | A4 | Steatosis_male | -0.289 | 0.132 | 0.036 | 0.166 | 35 | 14 |
| 11b-OHA4 | HOMAIR_male | 0.167 | 0.098 | 0.092 | 0.332 | 104 | 58 | B | Steatosis_male | -0.341 | 0.171 | 0.055 | 0.221 | 35 | 15 |
| E2 | HOMAIR_male | -0.161 | 0.096 | 0.095 | 0.332 | 104 | 50 | DOC | Steatosis_male | -0.318 | 0.163 | 0.060 | 0.224 | 35 | 19 |
| 11-KT | HOMAIR_male | 0.151 | 0.101 | 0.138 | 0.382 | 104 | 58 | 17a-OHP5 | Steatosis_male | -0.238 | 0.133 | 0.084 | 0.297 | 35 | 18 |
| AN | HOMAIR_male | -0.137 | 0.095 | 0.150 | 0.397 | 104 | 27 | E | Steatosis_male | -0.174 | 0.105 | 0.108 | 0.340 | 35 | 26 |
| E | HOMAIR_male | 0.130 | 0.100 | 0.196 | 0.475 | 104 | 62 | S | Steatosis_male | -0.253 | 0.185 | 0.180 | 0.457 | 35 | 20 |
| A | HOMAIR_male | 0.118 | 0.093 | 0.211 | 0.482 | 104 | 56 | 11-KT | Steatosis_male | -0.167 | 0.127 | 0.197 | 0.474 | 35 | 21 |
| E1 | HOMAIR_male | -0.119 | 0.093 | 0.207 | 0.482 | 104 | 50 | 17a-OHP4 | Steatosis_male | -0.137 | 0.110 | 0.223 | 0.514 | 35 | 27 |
| 17a-OHP5 | HOMAIR_male | -0.105 | 0.100 | 0.295 | 0.569 | 104 | 54 | 11-KDHT | Steatosis_male | -0.232 | 0.193 | 0.238 | 0.529 | 35 | 9 |
| 11-KDHT | HOMAIR_male | 0.087 | 0.097 | 0.372 | 0.650 | 104 | 20 | AN | Steatosis_male | -0.191 | 0.176 | 0.288 | 0.616 | 35 | 9 |
| DHEA | HOMAIR_male | -0.062 | 0.095 | 0.517 | 0.738 | 104 | 53 | 11b-OHA4 | Steatosis_male | -0.115 | 0.127 | 0.371 | 0.666 | 35 | 20 |
| P5 | HOMAIR_male | -0.066 | 0.101 | 0.512 | 0.738 | 104 | 50 | P4 | Steatosis_male | -0.049 | 0.060 | 0.427 | 0.674 | 35 | 4 |
| 17a-OHP4 | HOMAIR_male | -0.052 | 0.097 | 0.596 | 0.795 | 104 | 55 | DHT | Steatosis_male | -0.094 | 0.125 | 0.455 | 0.682 | 35 | 31 |
| DOC | HOMAIR_male | -0.051 | 0.102 | 0.622 | 0.795 | 104 | 52 | 11-KA4 | Steatosis_male | 0.139 | 0.213 | 0.519 | 0.692 | 35 | 32 |
| B | HOMAIR_male | -0.032 | 0.103 | 0.753 | 0.861 | 104 | 48 | T/Epi-T | Steatosis_male | -0.040 | 0.059 | 0.503 | 0.692 | 35 | 35 |
| A4 | HOMAIR_male | 0.007 | 0.097 | 0.945 | 0.996 | 104 | 56 | E2 | Steatosis_male | 0.019 | 0.056 | 0.735 | 0.852 | 35 | 13 |
| P4 | HOMAIR_male | 0.003 | 0.089 | 0.971 | 0.996 | 104 | 32 | A | Steatosis_male | -0.038 | 0.170 | 0.825 | 0.884 | 35 | 11 |
| 11-KA4 | HOMAIR_male | 0.000 | 0.101 | 0.998 | 0.998 | 104 | 94 | E1 | Steatosis_male | -0.025 | 0.107 | 0.817 | 0.884 | 35 | 10 |

## Notes

### Competing Interest Statement

The authors have declared no competing interest.

### Summary of Updates

Updated funding statement. Improved graphics in figures (no change in scientific content). Improved formatting in manuscript and supplement. There is no content change to text in the manuscript nor supplement.

